# Persistent organic pollutant concentrations in human pancreas and peripancreatic adipose tissues correlate with markers of beta cell dysfunction

**DOI:** 10.1101/2025.04.26.25326263

**Authors:** Myriam P. Hoyeck, Ma. Enrica Angela Ching, Jana Palaniyandi, Evgenia Fadzeyeva, Jelena Kolic, Jillian Ashley-Martin, James Lyon, Nancy Smith, Erin E. Mulvihill, James D. Johnson, Jocelyn E. Manning Fox, Patrick E. Macdonald, Jennifer E. Bruin

## Abstract

Epidemiological studies consistently report associations between circulating concentrations of persistent organic pollutants (POPs) and increased type 2 diabetes risk. Measures of POP concentrations in pancreas are limited; given the role of the endocrine pancreas in diabetes pathogenesis, this is an important gap in the literature. Additionally, no studies have correlated POP concentrations with direct measures of beta cell function in humans. We hypothesized that lipophilic POPs accumulate in human pancreas and correlate with markers of diabetes risk. To test this hypothesis, we measured POP concentration from 3 chemical classes – dioxins/furans, polychlorinated biphenyls (PCBs), and organochlorine pesticides (OCPs) – in pancreas and peripancreatic adipose tissue biopsies obtained from 31 human organ donors via the Alberta Diabetes Institute IsletCore. Indeed, POPs were consistently detected in human pancreas, and for some pollutants, at higher concentrations than in adipose. We next assessed correlations between POP concentrations and systemic indicators of diabetes risk (BMI, age, and %HbA1c) and direct measures of beta cell function. To this end, we measured insulin secretion in response to numerous secretagogues (i.e. glucose, fatty acids, amino acids, exendin-4, or KCl) in isolated islets from the same 31 donors. Pancreas PCBs and OCPs positively correlated with BMI, age and basal insulin secretion but negatively correlated with stimulation index (ratio of insulin secretion under high glucose / low glucose conditions). In contrast, pancreas dioxins/furans positively correlated with fatty acid- and amino acid-stimulated insulin secretion. These data confirm that lipophilic pollutants accumulate in human pancreas and positively correlate with markers of diabetes risk.

## 1. Introduction

Persistent organic pollutants (POPs) are a diverse family of environmental toxicants that resist degradation, leading to widespread bioaccumulation. POPs consist of a range of substances including intentionally produced chemicals (e.g., industrial chemicals and pesticides) and unintentionally produced chemicals (e.g., by-products of industrial processes); the most commonly encountered POPs in the environment include polychlorodibenzo-*p*-dioxins (PCDDs) and polychlorodibenzofurans (PCDFs), polychlorinated biphenyls (PCBs), and organochlorine pesticides (OCPs) [1,2]. The main source of human exposure to POPs is through consumption of contaminated foods (e.g., meat and dairy products) [3], although humans are also exposed through air particles [2]. Despite regulations to reduce the production and use of POPs, they remain detectable in our environment and in humans [4–6], making them a global concern for human health. Specifically, there is a large body of evidence – ranging from population-level studies to more detailed mechanistic investigations in rodent models – implicating POPs in type 2 diabetes (T2D) pathogenesis [7].

PCDDs/PCDFs, commonly referred to as dioxins and furans, are polyhalogenated POPs that are formed as by-products of combustion processes such as waste incineration and chemical manufacturing [8]. There are also natural sources of dioxins such as forest fires and volcanic eruptions. Dioxins/furans exert biological effects through activation of the aryl hydrocarbon receptor (AhR) pathway and AhR target genes, including the cytochrome P450 enzymes (*CYP1A1*/*1A2*). The most toxic and well-studied dioxin is 2,3,7,8-tetrachlorodibenzo-*p*-dioxin (TCDD); thus the toxicity of dioxins and dioxin-like chemicals is often expressed relative to TCDD [9,10]. Importantly, dioxins/furans have an estimated half-life of 7 – 11 years [11,12], and despite declining emissions over the past 20 years [13,14], some analytes remain detectable in >99% of the general Canadian population [15,16]. In fact, a recent cross-sectional study measured 7 dioxin and 10 furan analytes in plasma samples collected from a community in Yukon, Canada in 2022 (n = 54), and reported that 6 analytes were detected in >67% of serum samples, with 2 dioxin analytes having 100% detection rates [15]. Serum dioxin levels have been associated with increased T2D risk in the general population [7,17–20]; however, some studies also reported no associations between dioxins and T2D or HOMA-IR (homeostatic model assessment for insulin resistance) [21–24].

PCBs are synthetic chemicals that were widely used in commercial processes (e.g., dielectrics, heat exchange fluids, paint additives, and plastics) prior to being banned in the 1970s [25–27]. PCBs can be classified as “non-dioxin like” (NDL) or “dioxin-like” (DL) based on their affinity for AhR and mechanism of action that resembles dioxins [28]. The half-life of PCBs varies significantly depending on the congener and study population, with reports ranging from 0.34 – 300 years [29–31]. Several PCB analytes are detected in >90% of the Canadian population [15,16,32,33]. Specifically, a cross-sectional study on a Canadian Inuit population (n = 2172) measured 14 PCB analytes in plasma collected between 2007 – 2008, and reported detection rates >50% for 10 different PCBs, of which 6 analytes were detected in >90% of the sample population [32]. In the general population, serum PCB concentrations have been associated with prediabetes, HOMA-IR, abnormal glucose tolerance, and fasting hyperglycemia [7,17,18,21,34–36].

OCPs are a class of chlorinated organic compounds that were widely used as insecticides in agriculture and for the control of parasite-borne diseases [37]. OCPs of prominent concern include dichlorodiphenyltrichloroethane (DDT) and its metabolites dichlorodiphenyldichloroethylene (DDE) and dichlorodiphenyldichloroethane (DDD), hexachlorocyclohexane (HCH) and hexachlorobenzene (HCB) isomers, and chlordane [38]. Although many OCPs were banned or phased out in the USA and Canada in the 1970s – 1980s, they continue to be used in developing countries; DDT, HCH, aldrin and dieldrin are some of the most widely used pesticides in Asia [39,40]. As a result, Canadian are still exposed to OCPs through long-range atmospheric transport of these compounds, with some OCPs being detected in >90% of the general population [32,33]. For example, human biomonitoring data from the Canadian Health Measures Survey collected in 2007-2009 (n = 59) reported that of the 14 OCP analytes measured, 7 analytes had a detection rate of >50%, of which 4 analytes were measured in >90% of the general population. Serum OCP levels are consistently associated with increased risk of prediabetes and T2D, HOMA-IR, glucose intolerance, and fasting hyperglycemia [7,17,18,21,32,34–36,41–45].

Although there is mounting evidence that POPs are positively associated with increased risk of developing T2D and/or dysglycemia, these studies are largely based on serum POP levels since using serum is less invasive and more accessible than tissue biopsies. However, POPs are highly lipophilic compounds and preferentially accumulate in lipid-rich tissues such as adipose [46–48]; thus, serum concentrations can underestimate POP body burden [49,50]. We only found three studies that correlated POP levels in adipose tissue to T2D risk and all reported a positive association [51–53], although these studies only analysed a small subset of PCBs, OCPs, and fragrance substances. Despite not being traditionally considered a fatty tissue, it is well established that ectopic fat can accumulate in the pancreas [54,55], thus rendering the pancreas a potential target for POP accumulation. In fact, a limited number of studies measured POP levels—including dioxins/furans, PCBs, and 2 PFAS (per- and polyfluoroalkyl substances)— in human adipose and pancreas and reported comparable POP concentrations in both tissues [56,57]. However, no studies to date have assessed whether POP concentrations in pancreas tissue correlate with increased diabetes risk; given the role of the pancreas in metabolic disease, this is an important gap in the literature. Additionally, studies assessing whether POP concentrations (circulating or tissue-specific) correlate with direct markers of beta cell function in humans are lacking. Previous work has relied exclusively on fasting and glucose-induced plasma insulin levels as indirect measures of beta cell function, but plasma insulin is a poor proxy for beta cell function since it can be influenced by peripheral insulin action and insulin clearance [58].

The goal of the current study was to determine whether POP accumulation in human pancreas correlates with markers of diabetes risk, and whether these correlations are consistent with findings in adipose tissue. We measured dioxin/furan, PCB, and OCP concentrations (61 analytes total) in the pancreas and peripancreatic adipose tissue from 31 human organ donors (14 females, 17 males). We first assessed correlations between POP concentrations and systemic markers of diabetes/pre-diabetes risk, including age, BMI, and %HbA1c (% hemoglobin A1C; a measure of average blood glucose levels over the past 2 – 3 months and an indicator of diabetes status). Given that beta cell dysfunction precedes changes in %HbA1c [59,60], we also assessed insulin secretion in response to diverse secretagogues (glucose, oleate/palmitate, leucine, the glucagon like peptide 1 receptor (GLP1R) agonist exendin 4, and KCl) in isolated islets from the same 31 donors as a more direct and sensitive method of assessing diabetes risk. Our study provides unique evidence that pancreas POP concentrations directly correlate with measures of beta cell dysfunction in humans.

## 2. Materials and Methods

### 2.1. Human donor sample collection

Human donor tissues were processed at the Alberta Diabetes Institute (ADI) IsletCore (www.humanislets.com) [61] between 2019 – 2022 (see **Figure 1** for study design). A total of 31 human donors were analysed, including 14 females and 17 males. Donor ages ranged between 25 – 70, BMI ranged from 12.8 – 40.3, and %HbA1c ranged from 3.6 – 8.6 (**Figure 1A–C, Supp Table 1**); all donor characteristics are publicly available on the IsletCore website. All research using human islets was approved by the Research Ethics Board at Carleton University (#106701) and the University of Alberta (Pro00013094).

**Figure 1:**
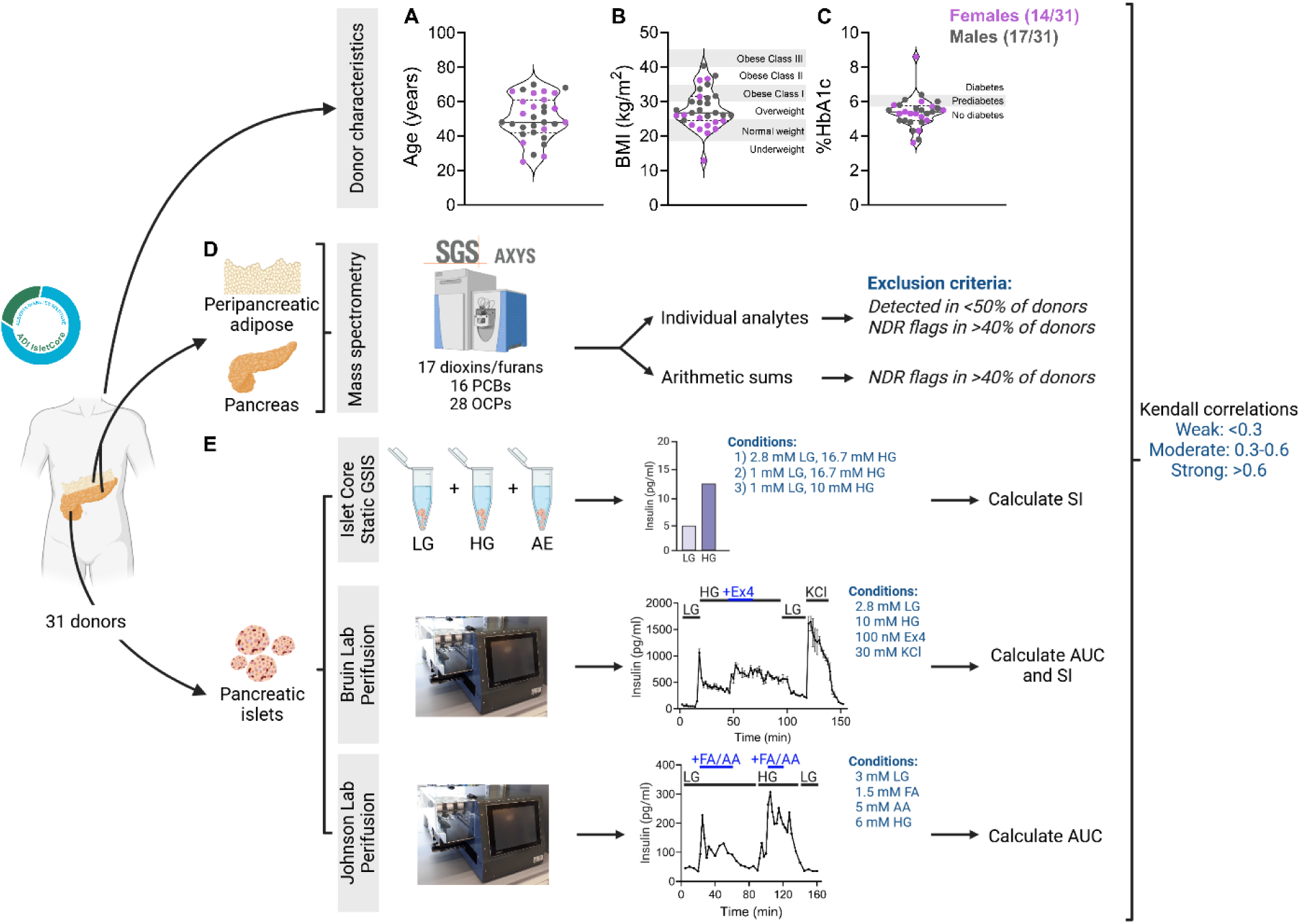
Experimental design schematic. Tissues from 31 human organ donor (14 females and 17 males) were obtained through the ADI IsletCore. **(A)** Age **(B)** BMI, and **(C)** % HbA1c is reported for all donors (female and male pooled). **(D)** Peripancreatic adipose and whole pancreas tissue were collected from each donor for mass spectrometry analysis to measure dioxin/furan, polychlorinated biphenyls (PCBs), and organochlorine pesticide (OCP) concentrations (pancreas n = 29, adipose n = 31 donors). We used both individual analyte concentrations and arithmetic sums when doing correlations. For individual analyte analysis, analytes that had NDR flags (i.e. peak detected but did not meet quantification criteria) in > 40% of donors and/or were detected in < 50% of the donors were excluded from the analysis. When doing arithmetic sums, analytes that had NDR flags in > 40% of donors were excluded. **(E)** Pancreatic islets were isolated from the same donors for functional assessments by three separate research labs. Static glucose-stimulate insulin secretion assays (GSIS) were conducted by the IsletCore using three combinations of glucose concentrations; stimulation index was calculated as a ratio of insulin concentration under high glucose (HG) relative to low glucose (LG) (n = 31 donors). Perifusion analysis was conducted by the Bruin lab, which included stimulation with 2.8 mM LG, 10mM HG, 10mM glucose + 100nM exendin4, and 30mM KCl; area under the curve and stimulation index were calculated for each donor (n = 30 donors). Perifusion analysis was conducted by the Johnson lab, which included stimulation with 3mM LG, 3mM LG + 1.5mM oleate/palmitate or 5mM leucine, 6 mM HG, and 6mM LG + 1.5mM oleate/palmitate or 5mM leucine; area under the curve was calculated for each donor (n = 23 donors). Kendall’s coefficient was used to assess correlations between pollutant concentrations and 1) pollutant concentrations within and across tissues, 2) donor characteristics, and 3) beta cell function.

We first attempted to measure POP levels in isolated islets from human donors (n = 3); a total of 4000 islets were hand-picked in a cryovial, washed 1x with PBS, and flash frozen. Unfortunately, POP detection rates were too low using this number of isolated islets (**Supp Fig 2**); as such we proceeded with intact pancreas biopsies for subsequent analysis.

**Figure 2:**
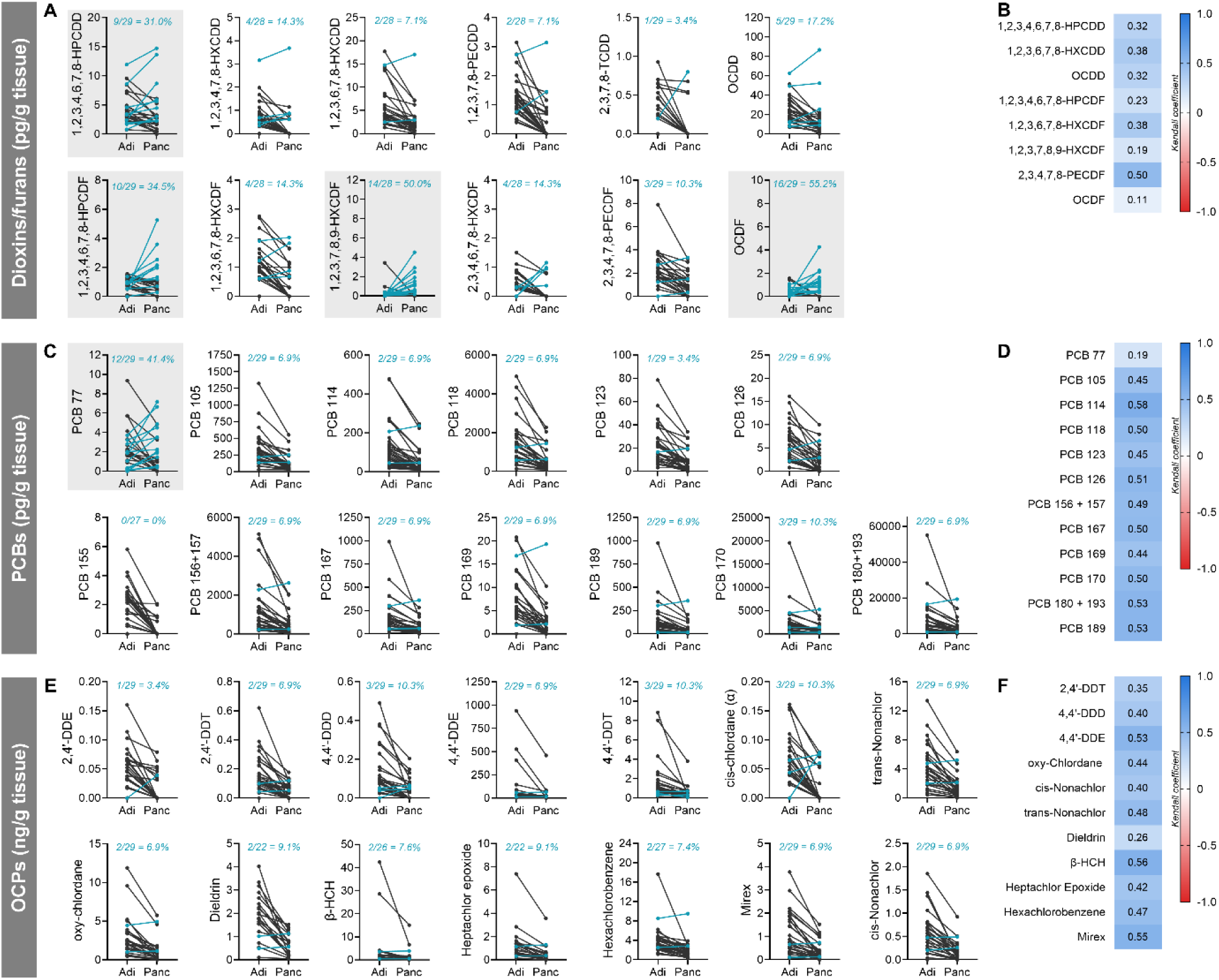
A subset of dioxin/furan and PCB analytes accumulate at higher concentrations in pancreas compared to adipose. **(A,C,E)** Before-after plots comparing **(A)** dioxin/furan, **(C)** polychlorinated biphenyl (PCB), and **(E)** organochlorine pesticide (OCP) concentrations in donor adipose (Adi) and pancreas (Panc) tissue. Adipose and pancreas concentrations from the same donor are connected by a line. Turquoise dots/lines represent donors that have higher analyte concentrations in pancreas compared to adipose; turquois text indicates the number and percentage of donors that have higher analyte concentrations in pancreas compared to adipose. Grey background emphasizes analytes that accumulate at higher concentrations in pancreas than adipose in >30% of donors. Analyte concentrations presented in before-after plots were blank corrected. **(B,D,F)** Heatmaps correlating **(B)** dioxin/furan, **(D)** PCB, and **(F)** OCP analyte concentrations in adipose and pancreas; only analytes with a detection rate > 50% in both tissue were retained for this analysis. Analyte concentrations used in the heatmaps were blank corrected and any zeroes were assigned a value of 1/2 LOD. Data in the heatmaps represent Kendall’s rank correlation coefficients.

**Figure 3:**
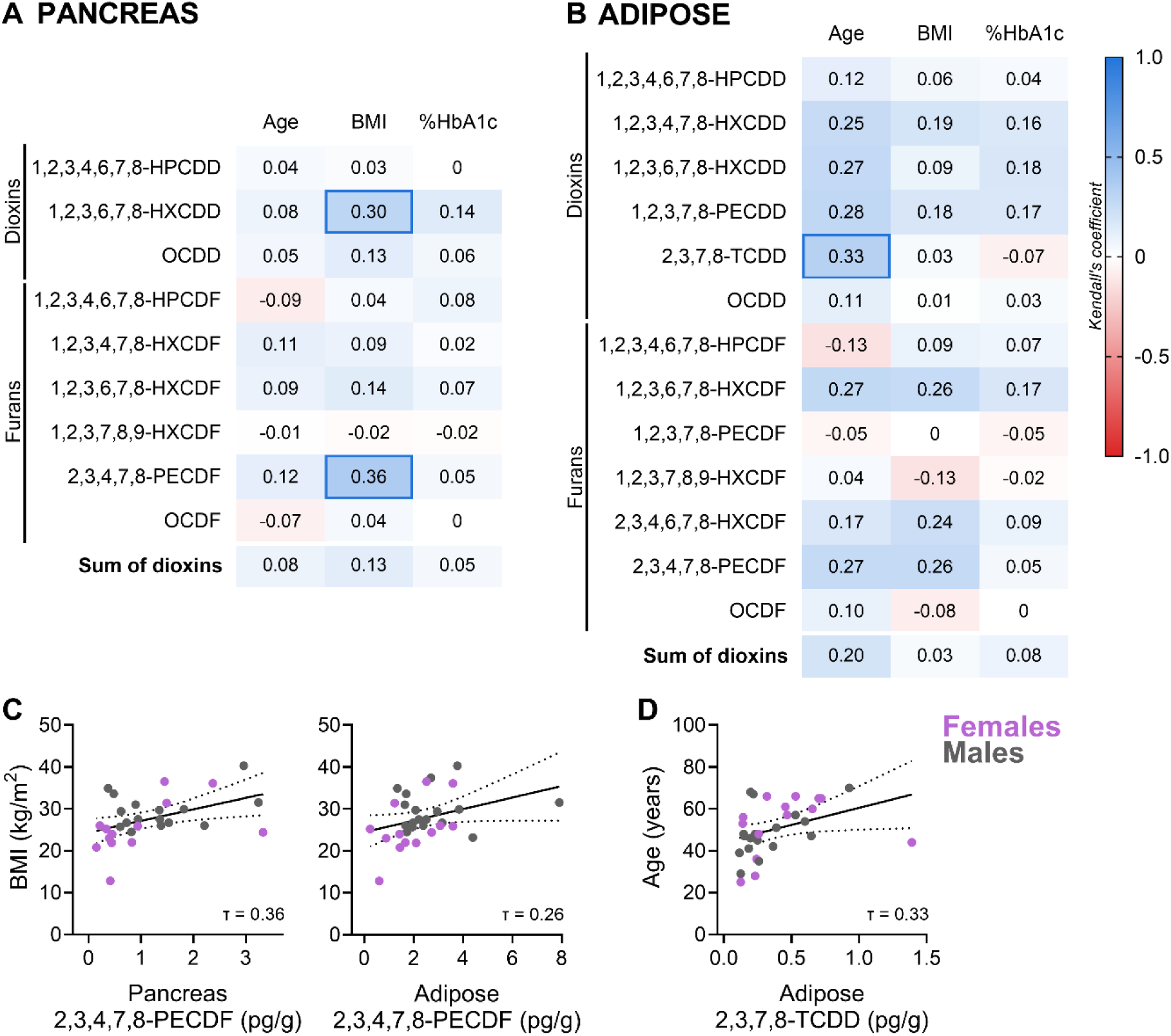
Dioxin/furan concentrations in both pancreas and adipose showed no overall correlations with donor characteristics. **(A,B)** Heatmaps depicting correlations between dioxin/furan concentrations in **(A)** pancreas and **(B)** adipose and donor characteristics, including age, BMI, and %HbA1C. **(C,D)** Scatter plots show individual donor correlations between **(C)** pancreas and adipose 2,3,4,7,8-PECDF and BMI, and **(D)** adipose 2,3,7,8-TCDD and age. Individual dots represent different donors, colour-coded by sex. All analyte concentrations used in this analysis were blank corrected and any zeros were assigned a value of 1/2 LOD. Data in the heatmaps and scatter plots represent Kendall’s rank correlation coefficients (τ).

For each donor, a ∼5 mm cubic biopsy of pancreas (0.077 – 0.442 g) and peripancreatic adipose (0.088 – 1.00 g) tissues were collected and flash frozen, with the exception of 2 donors that had an adipose biopsy but no pancreas biopsy (n = 31 adipose, n = 29 pancreas). The remaining pancreas tissue was then processed for pancreatic islet isolation, as described by the ADI IsletCore [62]. Isolated islets were cultured in CMRL media (Corning, #15110CV) supplemented with 0.5% BSA (30% v/v; Equitech Bio Inc, #BAL62), 1% insulin-transferrin-selenium (100x; Corning, #25800CR, Tewsbury, MA), 2% Gibco® GlutamMAX^TM^ (100x; ThermoFisher, #35050061, Waltham, MA), and 0.5% penicillin-streptomycin (Lonza, #09-757F).

### 2.2. Gas chromatography tandem mass spectrometry (GC-MS/MS)

The frozen donor pancreas and peripancreatic adipose biopsies were sent to SGS AXYS (Sidney, BC, Canada) to measure a panel of 17 dioxin/furan, 16 PCB, and 28 OCP analytes using GC-MS/MS (**Fig 1D**); AXYS is ISO 17025 accredited by the Canadian Association for Laboratory Accreditation. A list of all analytes measured can be found in **Supp Fig 4–6** and a summary of the OCP analytes and their degradation pathways is found in **Supp Fig 1**. Note that PCB 157 + 157 and PCB 180 + 193 were co-extracted, as such concentrations of these analytes could not be distinguished and are reported as a sum concentration for both analyte combinations.

**Figure 4:**
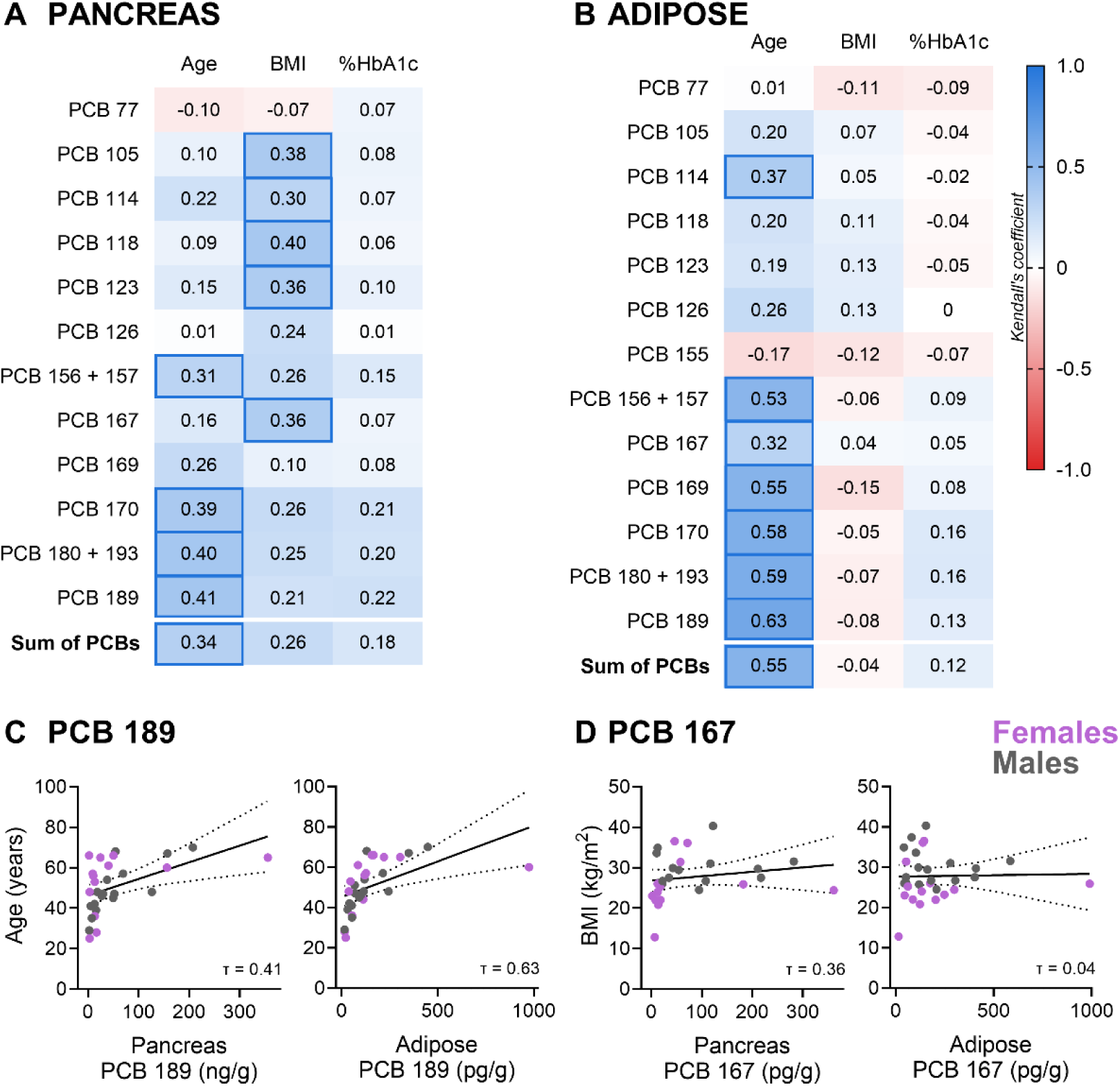
PCBs in pancreas positively correlate with age and BMI, whereas PCBs in adipose only positively correlated with age. **(A,B)** Heatmaps depicting correlations between polychlorinated biphenyl (PCB) concentrations in **(A)** pancreas and **(B)** adipose and donor characteristics, including age, BMI, and %HbA1C. **(C,D)** Scatter plots show individual donor correlations between **(C)** pancreas and adipose PCB 189 concentrations and age, and **(D)** pancreas and adipose PCB 167 concentrations and BMI. Individual dots represent different donors, colour-coded by sex. All analyte concentrations used in this analysis were blank corrected and any zeros were assigned a value of 1/2 LOD. Data in the heatmaps and scatter plots represent Kendall’s rank correlation coefficients (τ).

Dioxins/furans, PCBs, and OCPs were co-extracted from the same tissue biopsy. Pollutant concentrations in adipose and pancreas were analysed in 7 separate batches of 1 – 10 donors per batch; adipose and pancreas biopsies from a given donor were analysed in the same batch. All analysis batches included the samples, a procedural blank, and a spiked matrix (canola oil). In brief, all samples were homogenized and Soxhlet extracted with dichloromethane/hexane (1:1). After extraction, samples were spiked with cleanup standards and cleaned up using standard chromatographic columns. Extracts were subsequently fractionated for each analysis, spiked with ^13^C-labeled recovery standards and analysed using one of the following methods: dioxins/furans, MLA-217 as per EPA method ATM 16130; PCBs, MLA-210 using APGC-MS/MS; OCPs, MLA-228 using APGC-MS/MS. Final sample concentrations were determined based on labelled surrogate standard quantification and sample weight, and were adjusted for % recovery. Lab blanks and % analyte recovery per batch are summarized in **Supp Table 7**. Sample limits of detection (LOD) were established based on a 3:1 signal to background noise ratio, and were determined for each analyte and sample. The range of LOD values in our analysis are reported in **Supp Fig 4–6**.

Analytes that had NDR flags (i.e., defined by SGS AXYS as a peak that was detected but did not meet quantification criteria) in > 40% of donors were excluded from all analyses. This included 2 dioxin/furans in adipose, 1 dioxin/furan in pancreas, and 2 OCPs in both adipose and pancreas, leaving a total of 57 and 58 analytes in adipose and pancreas, respectively. Of the remaining analytes, those with a non-detect (ND) flag in > 50% of the donors were excluded from correlation analyses. As such, of the 17 dioxins/furans initially measured, 13 and 9 dioxin/furan analytes met all inclusion criteria in adipose and pancreas tissue, respectively (**Supp Fig 4A,C**). Of the 14 PCBs measured, 13 and 12 analytes were retained for analysis in adipose and pancreas, respectively (**Supp Fig 5A,C**). Of the 28 OCPs measured, 15 and 12 analytes were retained for analysis in adipose and pancreas, respectively (**Supp Fig 6A,C**).

**Figure 5:**
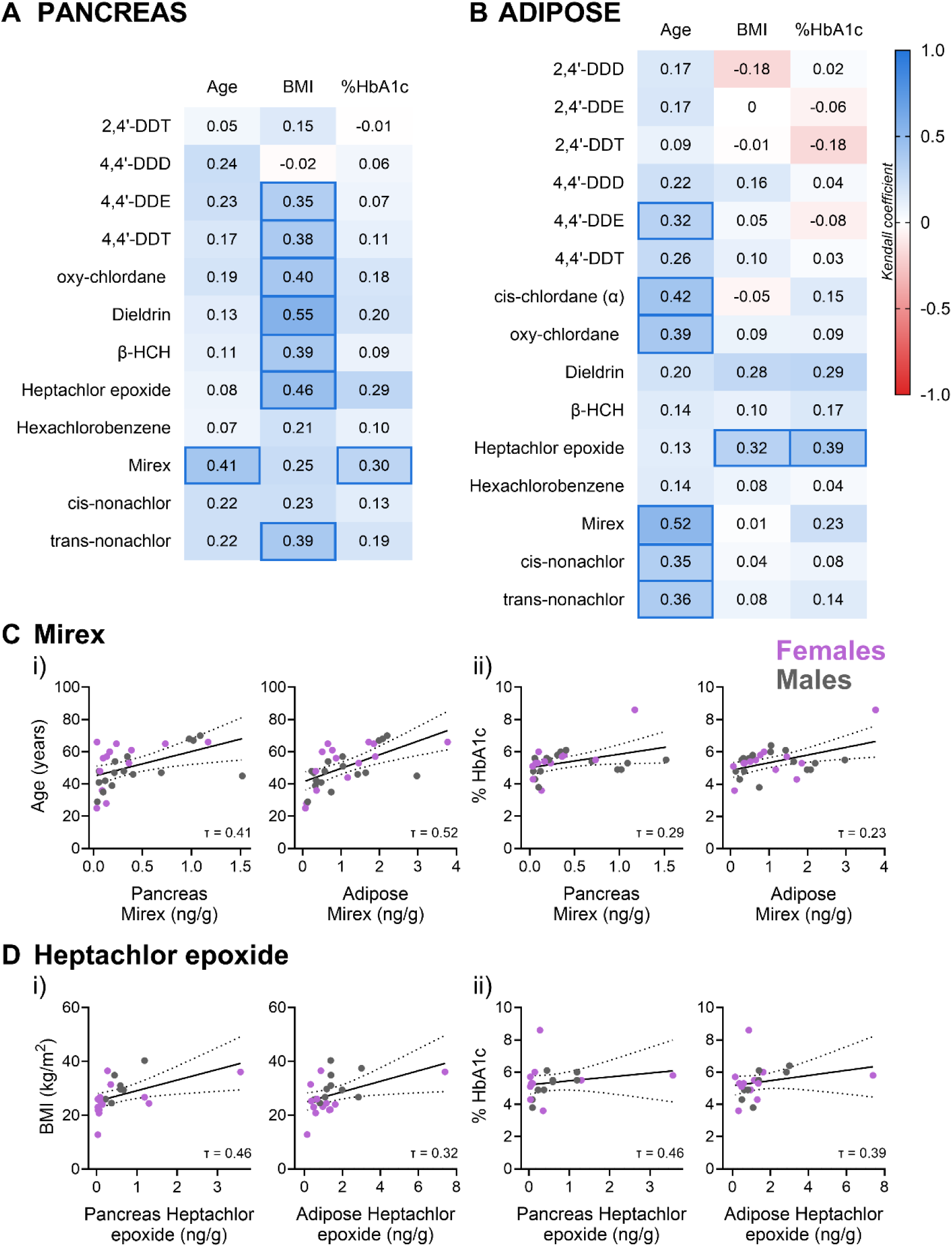
OCPs in pancreas positively correlate with BMI, whereas OCPs in adipose positively correlate with age. **(A,B)** Heatmaps depicting correlations between organochlorine pesticide (OCP) concentrations in **(A)** pancreas and **(B)** adipose and donor characteristics, including age, BMI, and %HbA1C. **(C,D)** Scatter plots show individual donor correlations between **(C)** pancreas and adipose mirex concentrations and (i) age and (ii) %HbA1c, and **(D)** pancreas and adipose heptachlor epoxide concentrations and (i) BMI and (ii) %HbA1c. Individual dots represent different donors, colour-coded by sex. All analyte concentrations used in this analysis were blank corrected and any zeros were assigned a value of 1/2 LOD. Data in the heatmaps and scatter plots represent Kendall’s rank correlation coefficients (τ).

**Figure 6:**
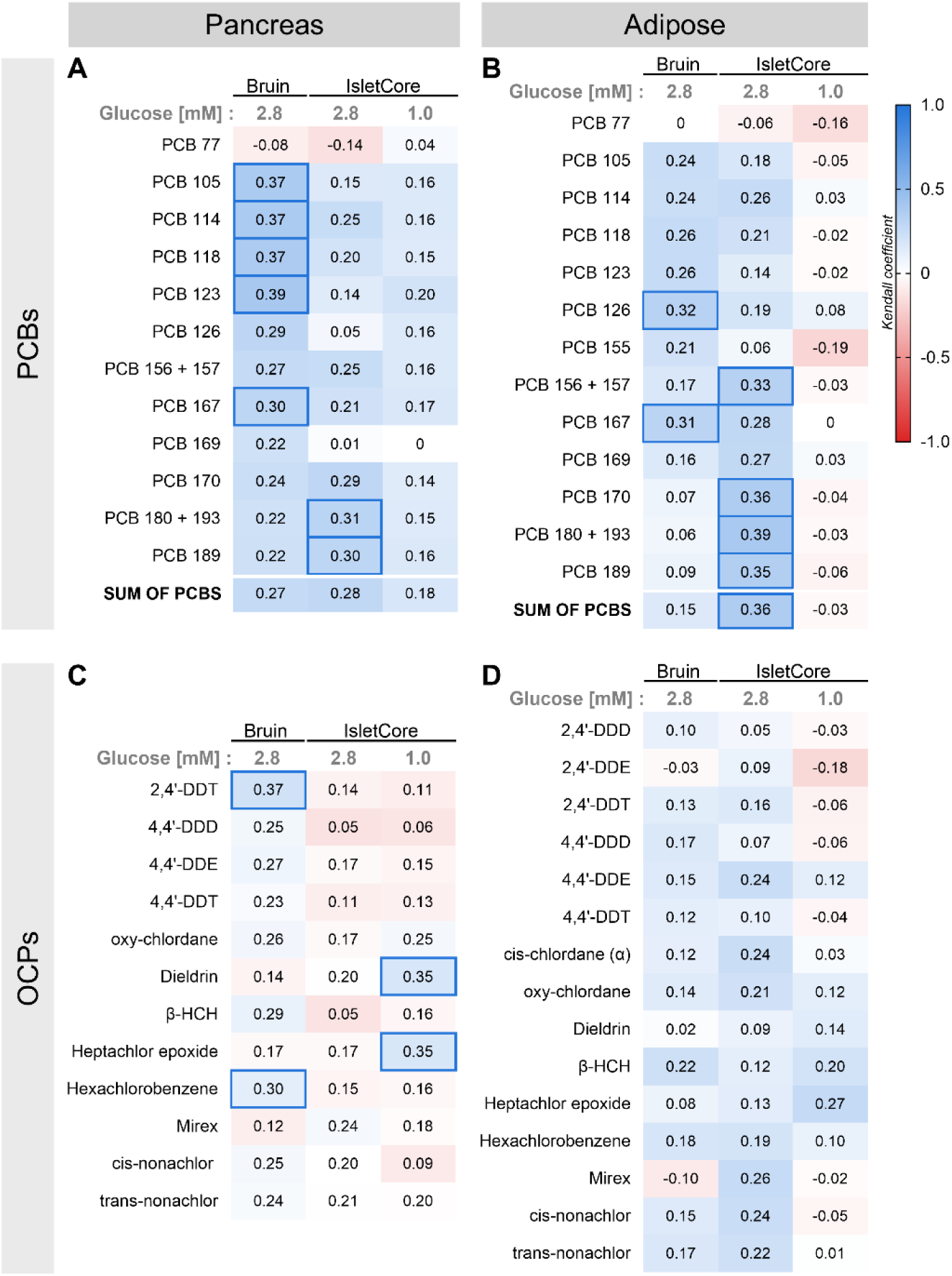
PCB, but not OCP, concentrations in pancreas and adipose were positively correlated with elevated insulin secretion under low glucose conditions. Heatmaps depicting correlations between **(A,B)** polychlorinated biphenyls (PCBs) and **(C,D)** organochlorine pesticides (OCPs) concentrations in human donor **(A,C)** pancreas and **(B,D)** adipose and insulin secretion under low glucose (LG) conditions. Human islets were stimulated with 1.0 or 2.8 mM low glucose, and total insulin secretion was measured following each stimulus. All analyte concentrations used in this analysis were blank corrected and any zeros were assigned a value of 1/2 LOD. Data in the heatmaps represent Kendall’s rank correlation coefficients. Bolded boxes emphasize correlations that were moderate to strong.

Analytes that passed both inclusion criteria were blank-corrected using batch-specific lab blank values (**Supp Fig 7**). For analytes that were retained for correlation analysis, any donors that had an ND flag for that analyte were assigned a value equivalent to half the LOD for that given donor.

**Figure 7:**
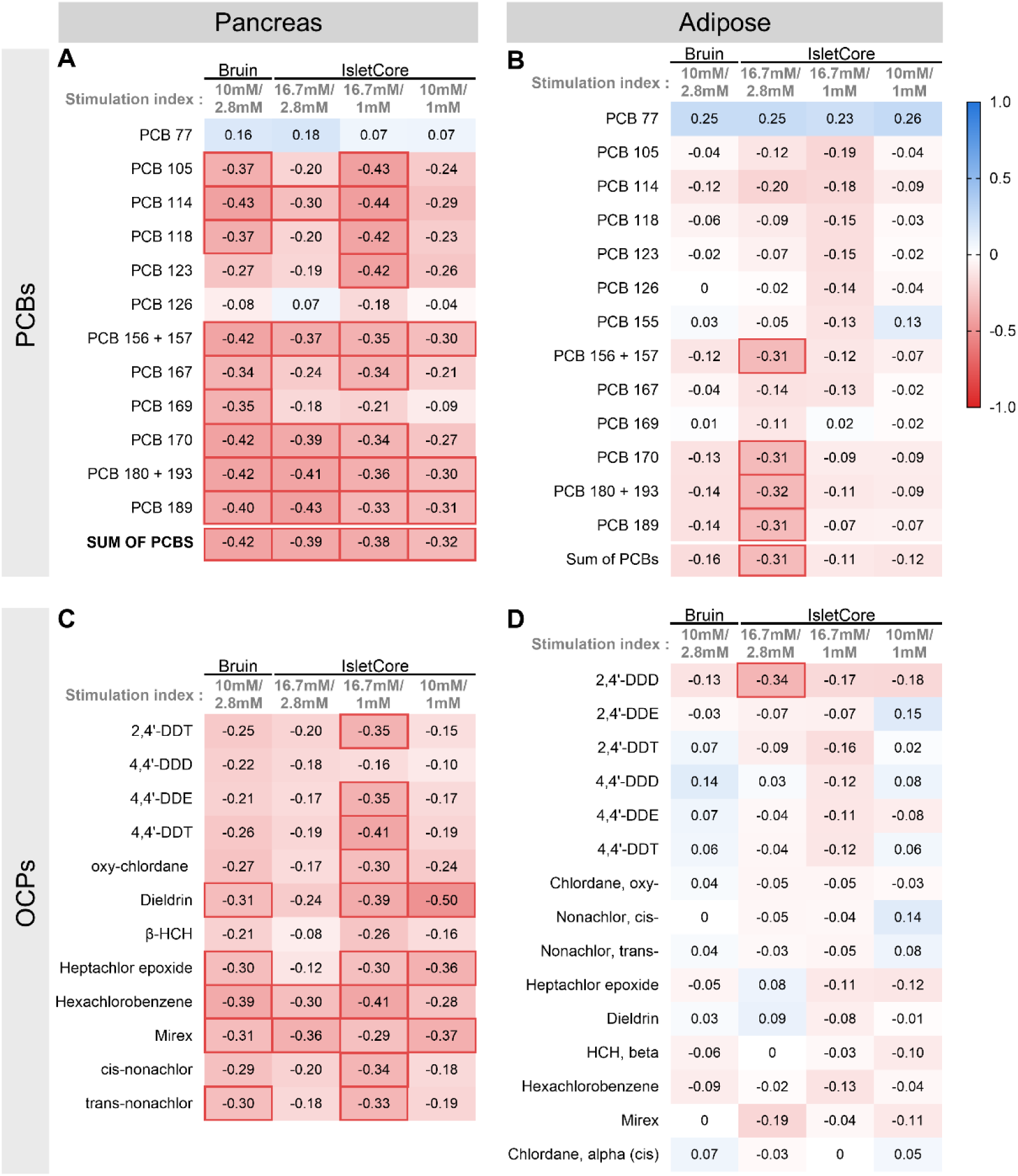
PCB and OCP concentrations in pancreas largely show negative correlations with stimulation index, with the exception of PCB 77 and PCB 126. Heatmaps depicting correlations between **(A,B)** polychlorinated biphenyls (PCBs) and **(C,D)** organochlorine pesticides (OCPs) concentrations in human donor **(A,C)** pancreas and **(B,D)** adipose and stimulation index. Human islets were stimulated with 1.0 or 2.8 mM low glucose (LG), followed by 10 or 16.7 mM high glucose (HG). Total insulin secretion was measured following each stimulus and stimulation index was calculated as a ratio of insulin concentration under HG relati ve to LG condition. All analyte concentrations used in this analysis were blank corrected and any zeros were assigned a value of 1/2 LOD. Data in the heatmaps represent Kendall’s rank correlation coefficients. Bolded boxes emphasize correlations that were moderate to strong.

### 2.3. Assessment of beta cell function

#### 2.3.1. Static glucose-stimulated insulin secretion analysis (IsletCore, University of Alberta)

A static glucose-stimulated insulin secretion (GSIS) assay was conducted at the ADI IsletCore for each donor (n = 31), as described (www.isletcore.ca; **Figure 1E**)[63]. In brief, 15 islets per replicate (n = 3 technical replicates per donor) were immersed in low glucose (LG) and high glucose (HG) KRBB (Krebs– Ringer bicarbonate buffer; 115 mM NaCl, 5 mM KCl, 24 mM NaHCO3, 2.5 mM CaCl2, 1 mM MgCl2, 10 mM HEPES, 0.1% (wt/vol.) BSA, pH 7.4) for sequential 1-hour incubations with differing combinations of glucose concentrations, including: 1 mM LG followed by 10 mM HG, 1 mM LG followed by 16.7 mM HG, or 2.8 mM LG followed by 16.7 mM HG. Secreted insulin concentrations were measured in supernatant by Chemiluminescence ELISA (ALPCO, #80-INSHU-CH01; Salem, NH, USA). Technical replicates were averaged per donor. Stimulation index (SI) was calculated as the ratio of insulin concentration under HG relative to LG condition. Individual donor data are presented in **Supp Fig 8A–C**.

**Figure 8:**
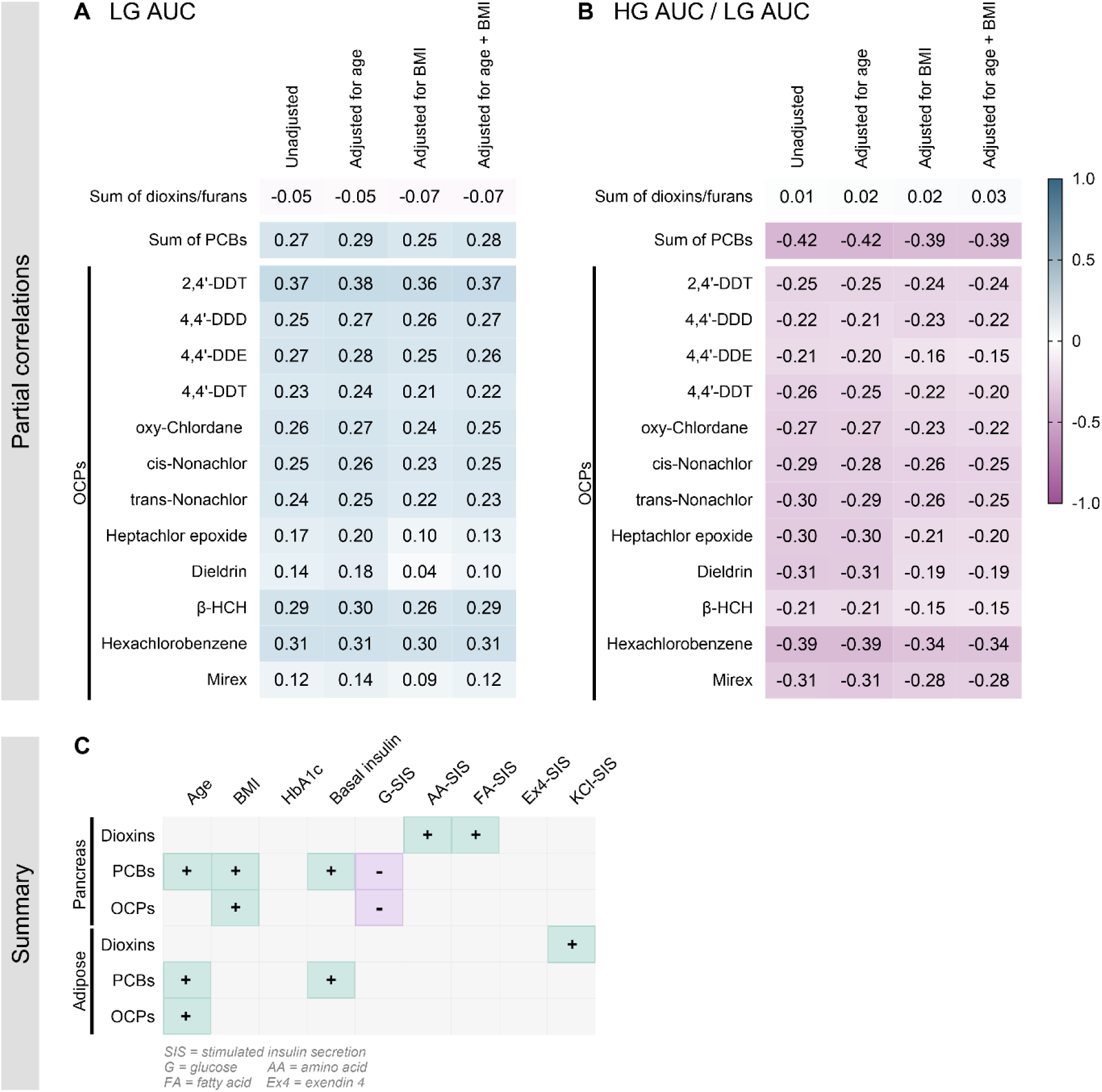
Associations between pancreas pollutant concentrations and altered insulin secretion were not changed when correcting for age and/or BMI. **(A,B)** Heatmaps displaying adjusted and unadjusted correlations between the sum of dioxin/furans, the sum of polychlorinated biphenyls (PCBs), and organochlorine pesticides (OCPs) in pancreas and **(A)** insulin secretion under low glucose conditions (LG AUC), and **(B)** stimulation index (HG AUC / LG AUC) using perifusion analysis from the Bruin lab. Data in the heatmaps represent Kendall’s rank correlation coefficients (τ) that were unadjusted or partial coefficients adjusted for age, BMI, or age + BMI. All analyte concentrations used in this analysis were blank corrected and any zeros were assigned a value of 1/2 LOD. **(C)** Graphical table summarizing the association between dioxin/furan, PCB, and OCP concentrations in pancreas and adipose with both donor characteristics (age, BMI, %HbA1c) and insulin secretion in response to various secretagogues (glucose, the amino acid leucine, the fatty acids oleate/palmitate, exendin4 (Ex4), and KCl). A “+” symbol with green shading indicates a positive correlation, whereas a “-“ symbol with purple shading indicates a negative association. Grey shading indicates no significant correlations.

#### 2.3.2. Dynamic perifusion analysis (Bruin Lab, Carleton University)

Islets were shipped in complete CMRL media overnight from Edmonton to Ottawa for functional assessments in the Bruin lab. Upon arrival in the Bruin lab, islets were transferred to DMEM media (Gibco, #11885-084; Waltham, MA, USA) supplemented with 10% fetal bovine serum (Sigma, #F1051; St. Louis, MO, USA) and 1% penicillin-streptomycin (Gibco, 15140-122-100), and cultured overnight at 37°C and 5% CO2 prior to conducting analyses to allow for recovery from shipment.

Dynamic insulin secretion was assessed in isolated human islets by perifusion (PERI4-115/230, Biorep Technologies; Miami, FL, USA; **Figure 1E**) in 30 of our 31 human donors. Perifusion was conducted 1 – 2 days after cells were received from the IsletCore. For each donor, 100 islets were handpicked per technical rep (n = 1 – 2 technical replicates per donor) and loaded in Perspex microcolumns between two layers of acrylamide-based microbeads (PERI-BEADS-20, Biorep Technologies). Islet were perfused for 48 min at a rate of 100 μL/min with KRBB containing 2.8 mM LG to equilibrate the islets (samples dscarded). Islets were then perfused with KRBB containing 2.8 mM LG for 12 min, 10 mM HG for 32 min, 10 mM HG + 0.1 μM Ex4 (Abcam, #ab120214; Cambridge, United Kingdoms) for 24 min, 10 mM HG for 24 min, 2.8 mM LG for 24 min, 30 mM KCl for 20 min, and 2.8 mM LG for 16 min. Samples were collected every 2 min (200 μl total per sample). Islets and perifusion solutions were kept at 37°C throughout the perifusion run using the built-in temperature-controlled chamber, and the collection plate was kept at 4°C using the built-in tray cooling system. Samples were kept at −80 °C for long-term storage. Insulin concentrations were measured by ELISA (ALPCO, #80-INSHU-CH01).

Individual perifusion curves are presented in **Supp Fig 8D–F**; technical replicates were averaged per donor. For correlation analysis, area under the curves (AUCs) were calculated for each perifusion stimulation step using the following analysis windows: LG AUC from 0 – 12 minutes, HG AUC from 14 – 44 minutes, HG + Ex4 AUC from 46 – 68 minutes, and KCl AUC from 118 – 136 minutes. Stimulation index following the HG stimulus was calculated as a ratio of total insulin concertation under HG relative to LG condition (i.e. HG AUC / LG AUC). Stimulation index following the KCl stimulus was calculated as a ratio of total insulin concertation under KCl relative to LG condition (i.e. KCl AUC / LG AUC). Due to technical reasons, some donor AUCs were excluded from analysis (summarized in **Supp Table 2**)

#### 2.3.3. Dynamic perifusion analysis (Johnson Lab, UBC)

A second perifusion analysis was conducted separately in the laboratory of Dr. James Johnson (UBC; Vancouver, BC) to assess nutrient-stimulated insulin secretion in a subset of the islet donors (n = 23; **Figure 1B**), as described in [64], and data was kindly provided to us. In brief, islets were perfused with KRBB containing 3 mM LG for 20 min, 3 mM LG + 1.5 mM oleate/palmitate (1:1 mix) for 40 min, 3 mM LG for 30 min, 6 mM HG for 10 min, 6 mM HG + 1.5 mM oleate/palmitate for 20 min, 6 mM HG for 10 min, and 3 mM LG for 30 min. A second subset of islets was perfused with KRBB containing 3 mM LG for 20 min, 3 mM LG + 5 mM leucine for 40 min, 3 mM LG for 30 min, 6 mM HG for 10 min, 6 mM HG + 5 mM leucine for 20 min, 6 mM HG for 10 min, and 3 mM LG for 30 min. Insulin concentrations were measured using a human insulin radioimmunoassay kit (Millipore, #HI-14K; Massachusetts, USA).

Individual perifusion data are presented in **Supp Fig 8G–I**. For correlation analysis, AUCs were calculated for each perifusion stimulation step using the following analysis windows: 3 mM LG + 5 mM leucine or 1.5 mM oleate/palmitate AUC from 25 – 60 min, and 6 mM HG + 5 mM leucine or 1.5 mM oleate/palmitate AUC from 105 – 120 min.

### 2.4. Statistical analysis and data visualization

#### 2.4.1. Kendall correlations

We used Kendall’s rank correlation coefficient (τ), a non-parametric test, to ensure robustness against outliers and suitability for small sample sizes [65]. We calculated correlations among: (1) analyte concentrations within the same pollutant class and tissue (e.g., dioxins vs. dioxins, PCBs vs. PCBs, and OCPs vs. OCPs), (2) analyte concentrations by pollutant class in pancreas vs. adipose (e.g., pancreas PCB vs. adipose PCB), (3) analyte concentrations and donor characteristics (e.g., age, BMI, Hba1c), and (4) analyte concentrations and indicators of beta cell function from static GSIS and dynamic perifusion assays. Unfortunately, our samples size was too small to stratify our data by sex and perform statistical tests. Instead, we visualized key correlations using scatter plots and colour coded the data points by sex to give a general representation of the distribution of our data.

To account for potential confounding effects of age and BMI on our results, we performed partial Kendall correlations adjusting for age, BMI, or both; age was self-reported while body mass index (BMI) was calculated using weight and height by the organ procurement organization. We computed correlation coefficients based only on complete cases for each pair of variables and used pairwise deletion to handle missing values. We categorized the strength of correlation as “weak” (τ < 0.3), “moderate” (0.3 ≤ τ ≤ 0.6), and “strong” (τ > 0.6) [65]. Consistent with the guidance from the American Statistical Association, we interpret our coefficients based on strength of correlation as opposed to null hypothesis testing and p-values [66,67].

#### 2.4.2. Principal component analysis and sensitivity analysis

We performed separate principal components analyses (PCA) for pancreas analytes and adipose analytes. Each analysis included pollutant concentrations for all dioxins/furans, PCBs, and OCPs within each tissue. We identified 3 donors in the pancreas dataset (R389, R419, R448) and 2 donors in the adipose dataset (R419, R421) who did not cluster with the remaining 27 donors in pancreas and 29 donors in adipose (**Supp Fig 3**), suggesting that these donors exhibit a different pollutant concentration profile than most donors. However, due to limitations in data availability, we cannot determine whether these differences are driven by environmental exposures or lifestyle factors. As such, we conducted sensitivity analyses on these donors to determine their impact on our overall results. We re-calculated Kendall’s correlation coefficients for all comparisons after systematically removing each donor individually and all identified donors simultaneously. Following these tests, we found that the overall impact of these donors was negligible (0.00 to 0.19 change in correlation strength); therefore, we kept them in our final analyses.

All statistical analyses were performed with R Statistical Software (version 4.4.2) [68]. For PCA analyses, we used *pcaMethods* (Bioconductor, version 1.98.0); we used the NIPALS imputation algorithm to address missingness [69]. All figures were made in GraphPad Prism 9.5.1.

## 3. Results

### 3.1. Dioxins/furans, PCBs, and OCPs are consistently detected in human donor adipose and pancreas

Once we excluded analytes due to NDR flags – leaving 57 and 58 analytes in adipose and pancreas, respectively – 41 analytes (72%) were detected in peripancreatic adipose tissue from >50% of our donor samples (**Supp** Fig 4**–6**). We also found consistent POP levels in pancreas, with 33 of the 58 analytes (57%) being detected in >50% of donor samples (**Supp** Fig 4**–6**).

Adipose and pancreas analyte concentrations ranged from 0.01 – 63.01 pg/g tissue and 0.14 – 86.58 pg/g tissue for dioxins/furans, 0.03 – 55,096.51 pg/g tissue and 0.29 – 19,392.36 pg/g tissue for PCBs, and 0.001 – 939.87 ng/g tissue and 0.001 – 458.87 ng/g tissue for OCPs, respectively (**Supp Fig Table 4 –6**). These data clearly demonstrate that POPs are accumulating in both human adipose and pancreas. OCPs are the most concentrated, followed by PCBs, and lastly dioxins/furans (**Supp Fig Table 4 –6**).

The most highly detected dioxin/furan analytes in adipose were OCDD > 1,2,3,6,7,8-HXCDD > 1,2,3,4,6,7,8-HPCDD > 2,3,4,7,8-PECDF (**Supp Fig 4A,B**), whereas in pancreas it was OCDD > 1,2,3,4,6,7,8-HPCDD > 1,2,3,6,7,8-HXCDD > 2,3,4,7,8-PECDF (**Supp Fig 4C,D**). Overall, dioxins were detected at higher concentration than furans (**Supp** Fig 4). Of note, the median TCDD concentration in adipose was 0.26 pg/g tissue and TCDD was detected in adipose from 58% of donors (**Supp Fig 4A,B**), whereas TCDD was detectable in pancreas from only 14% of donors (**Supp Fig 4C**). The most concentrated PCB analytes in adipose were PCB 180 + 193 > PCB 170 > PCB 118 > PCB 156 + 157 (**Supp Fig 5A,B**), whereas in pancreas it was PCB 180 + 193 > PCB 118 > PCB 170 > PCB 156 + 157 (**Supp Fig 5C,D**). Interestingly, PCB 180 + 193 had a concentration >2x greater than any other PCB analyte in both tissues (**Supp** Fig 5). The most concentrated OCP analytes in both adipose and pancreas were 4,4’-DDE > hexachlorobenzene > trans-nonachlor > oxy-chlordane > dieldrin > β-HCH (**Supp** Fig 6). 4,4’-DDE had a concentration >9x greater than any other OCP analyte in both tissues (**Supp** Fig 6).

Analytes from each class of pollutants largely showed moderate to strong positive correlations with each other within a tissue (**Supp** Fig 9**–11**), with the exception of PCB 77 in pancreas, which was negatively correlated with other PCB analytes (т = −0.35 to 0.07; **Supp Fig 10B**). These data suggest that analytes within a class of pollutants have similar patterns of accumulation in a tissue.

Collectively, these data indicate that legacy POPs that have been tightly regulated or banned are still detected in tissues from Canadian organ donors. We also show a diverse range of analyte concentration within a class of pollutants and across analytes classes, suggesting higher exposure rates or slower elimination of certain analytes; further analysis is needed to better understand exposure sources and the toxicokinetic of these POP analytes.

### 3.2. A subset of dioxin/furan and PCB analytes accumulate at higher concentrations in pancreas compared to adipose

Generally, POP concentrations were higher in adipose tissue compared to pancreas (Fig 2, **Supp** Fig 4**–6**). We also show that analyte concentrations between adipose and pancreas are positively correlated (**Fig 2B,D,F**), indicating accumulation of analytes across tissues. However, we identified a subset of analytes that accumulate at higher levels in pancreas compared to adipose, including 4 dioxins/furans (1,2,3,4,6,7,8-HPCDD, 1,2,3,4,6,7,8-HPCDF, 1,2,3,7,8,9-HXCDF, OCDF) and PCB 77 (**Fig 2A,C** – highlighted in grey). Of note, PCB 77 concentrations were inversely proportional to other PCB analyte concentrations in pancreas (**Supp Fig 10B**), suggesting that PCB 77 has a different pattern of accumulation in pancreas compared to all other PCBs. On average, OCP analytes were more concentrated in adipose compared to pancreas (**Fig 2E, Supp** Fig 6). These data indicate that some pollutants may preferentially accumulate in the pancreas rather than other fatty tissues such as adipose, and may point to some chemicals as being of particular concern for islet health/function.

### 3.3. Dioxin/furan concentrations in both pancreas and adipose showed no overall correlations with donor characteristics

We next assessed whether pollutant concentrations in human pancreas and adipose tissues correlated with clinical risk factors for diabetes. Two of the most abundant dioxins/furans in pancreas (1,2,3,6,7-HXCDD and 2,3,4,7,8-PECDF; **Supp** Fig 4) showed a moderate positive correlation with BMI (т = 0.30 and 0.36; **Fig 3A,C**), whereas these same analytes in adipose only show a weak positive to no correlation with BMI (т = 0.09 and 0.26; **Fig 3B,C**). Notably, TCDD concentrations in adipose showed a moderate positive correlation with age (т = 0.33; **Fig 3B,D**). Surprisingly, none of the dioxins/furans that preferentially accumulated in pancreas compared to adipose (**Fig 2A**) correlated with donor characteristics (Fig 3). There were no notable correlations between the sum of dioxin/furans in either pancreas or adipose and age, BMI, or %HbA1c (**Fig 3A,B**).

### 3.4. PCBs in pancreas positively correlate with age and BMI, whereas PCBs in adipose only positively correlated with age

Pancreas PCBs largely showed a moderate positive correlation with age and BMI (**Fig 4A,C,D**), whereas adipose PCB concentrations only correlated with age (**Fig 4B,C,D**). For example, pancreas and adipose PCB 189 had a correlation coefficient of 0.41 and 0.63 with donor age, respectively (**Fig 4C**), whereas pancreas and adipose PCB 167 had a correlation coefficient of 0.36 and 0.04 with BMI, respectively (**Fig 4D**). As expected, pancreas PCB 77 differed from other PCBs and showed a weak negative correlation with age and BMI (**Fig 4A**). There were no correlations between PCBs in either tissue and %HbA1c (**Fig 4A,B**).

### 3.5. OCPs in pancreas positively correlate with BMI, whereas OCPs in adipose positively correlate with age

Pancreas OCPs largely showed a moderate positive correlation with BMI (т = −0.02 to 0.55; **Fig 5A**), whereas adipose OCPs positively correlated with age (т = 0.09 to 0.52; **Fig 5B**). We identified an interesting subset of OCPs that correlated differently than the bulk of OCPs. Pancreas Mirex concentrations showed a moderate positive correlation with both age (т = 0.41; **Fig 5A,Ci**) and %HbA1c (т = 0.3; **Fig 5A,Cii**), and only a weak correlation with BMI (т = 0.25; **Fig 5A**). Adipose heptachlor epoxide (**Fig 5B,D**) and dieldrin (**Fig 5B**) concentrations showed a moderate to weak positive correlation with BMI and %HbA1c. These data point to a potential link between Mirex, heptachlor epoxide, and dieldrin, with increased hyperglycemia.

Collectively, our correlation analyses with donor characteristics suggest an association between pancreas POP concentrations and increased BMI, pointing to a link with peripheral tissue lipid accumulation. Adipose PCBs and OCPs generally showed a positive correlation with age. Interestingly, several pancreas PCBs also positively correlated with age, indicating that some POPs accumulate in pancreas over time. Lastly, we identified a subset of OCPs that might be linked to impaired glucose homeostasis.

### 3.6. PCB, but not dioxin/furan or OCP, concentrations in pancreas and adipose were positively correlated with elevated insulin secretion under low glucose conditions

It is well documented that elevated fasting insulin levels can precede the onset of T2D [70–72]. As such, we examined the relationship between POP concentrations and basal insulin secretion in isolated donor islets using perifusion data from our lab and static GSIS data from the ADI IsletCore. We did not observe any correlations between pancreas or adipose dioxins/furans concentrations and insulin secretion under low glucose conditions (**Supp Fig 12A,B**). Two OCP analytes (2,4’-DDT and hexachlorobenzene) in pancreas had a moderate positive correlation with basal insulin secretion from our perifusion analysis (**Fig 6C**), but these correlations were not reproduced using the IsletCore data. There were no other notable correlations between OCP concentrations and insulin secretion under low glucose conditions (**Fig 6C,D**).

Interestingly, pancreas and adipose PCB concentrations consistently showed a positive correlation with insulin secretion under 2.8 mM low glucose conditions from both our lab and the IsletCore (pancreas PCBsum т = 0.27 to 0.28; adipose PCBsum т = 0.15 to 0.36); the strength of these correlations was diminished when using a glucose concentration of 1.0 mM (**Fig 6A,B**). PCB 77 had a different trend and instead showed a weak negative correlation with insulin secretion under 2.8 mM low glucose conditions (Bruin т = −0.08, IsletCore т = −0.13; **Fig 6A,B**). These data suggest that PCB accumulation in both pancreas and adipose tissue is associated with increased insulin secretion under basal glucose conditions (i.e., representative of fasting glucose levels).

### 3.7. PCB and OCP concentrations in pancreas largely show negative correlations with stimulation index, with the exception of PCB 77 and PCB 126

Next, we investigated whether POP concentrations correlate with GSIS in isolated islets using dynamic perifusion from our lab and static GSIS data from the ADI IsletCore. Pancreas and adipose dioxins/furans did not correlate with insulin secretion under high glucose conditions (**Supp Fig 12C,D**) or with stimulation index (i.e., high glucose / low glucose ratio; **Supp Fig 12E,F**). Pancreas and adipose PCBs and OCPs did not show any notable correlations with total insulin secretion under high glucose conditions (**Supp** Fig 13).

Pancreas PCBs (PCBsum т= −0.42 to −0.32; **Fig 7A**) and OCPs (т= −0.41 to −0.08; **Fig 7C**) consistently showed a negative correlation with stimulation index, irrespective of the glucose concentration used to stimulation insulin secretion (10 or 16.7 mM). In contrast, only a subset of adipose PCBs (**Fig 7B**) and OCPs (**Fig 7D**) were negatively correlated with stimulation index, and only when a high glucose dose of 16.7 mM and a low glucose dose of 2.8 mM were used. Interestingly, PCB 77 and PCB 126 differed from other PCBs; pancreas and adipose PCB 77 had a weak positive correlation with stimulation index, whereas pancreas PCB 126 showed no overall correlation with stimulation index (**Fig 7A,B**).

In summary, the majority of PCB and OCP analytes in pancreas are associated with hypersecretion of insulin under basal glucose conditions and/or impaired stimulation index, whereas dioxin/furan are largely not correlated with changes in GSIS. PCB 77 and 126 had a different correlation pattern, suggesting that not all POP analytes affect GSIS in the same manner. Importantly, correlations between PCBs/OCPs and measures of insulin secretion were maintained when we ran partial correlation analysis and adjusted for age and/or BMI (**Fig 8A,B**), implying that correlations between POP analytes and donor characteristics are not driving correlations between POP analytes and beta cell function.

### 3.8. A subset of POPs in pancreas and adipose correlated with fatty acid-, amino acid-, Ex4-, and/or KCl-stimulated insulin secretion

Lastly, we assessed correlations between pollutant concentrations and insulin secretion in response to various other secretagogues that are known to amplify insulin secretion (fatty acids, leucine, exendin4), as well as KCl to assess insulin secretion capacity. Despite not correlating with insulin secretion following a HG stimulation, pancreas dioxins/furans showed an overall weak to moderate correlation with oleate/palmitate- and leucine-stimulated insulin secretion (**Supp Fig 14A**); no correlations were observed with Ex4- or KCl-stimulated insulin secretion (**Supp Fig 14A**). There were no overall correlations between the sum of adipose dioxins/furans and oleate/palmitate, leucine, or Ex4-stimulated insulin secretion, but a moderate positive correlation was seen with KCl-stimulated insulin secretion (**Supp Fig 14B**).

Only a subset of PCBs and OCPs showed correlations with oleate/palmitate, leucine, Ex4, or KCl-stimulated insulin secretion (**Supp Fig 14C–F**); notably, these are largely analytes that showed distinct correlations with donor characteristics (Fig 4**,5**) and/or GSIS (Fig 6**,7**). For example, pancreas PCB 77 positively correlated with both oleate/palmitate- and leucine-stimulated insulin secretion (т = 0.30 to 0.36; **Supp Fig 14C**). Pancreas PCB 126 positively correlated with leucine-stimulated insulin secretion, but only when co-stimulated with 6 mM glucose (т = 0.30; **Supp Fig 14C**), with Ex4-stimulated insulin secretion (т = 0.32; **Supp Fig 14C**), and with KCl-stimulated insulin secretion (т = 0.42; **Supp Fig 14C**); adipose PCB 126 positively correlated with both Ex4-(т = 0.31) and KCl-(т = 0.32) stimulated insulin secretion (**Supp Fig 14D**). Pancreas heptachlor epoxide positively correlated with leucine-stimulated insulin secretion (т = 0.36; **Supp Fig 14E**), whereas both adipose heptachlor epoxide (т = 0.41 to 0.66) and dieldrin (т = 0.25 to 0.50) positively correlated with oleate/palmitate and leucine-stimulated insulin secretion (**Supp Fig 14F**). These data suggest that pancreas dioxins/furans disrupt the insulin amplification pathway in response to macronutrients such as fatty acids and amino acids. Similarly, a small subset PCB and OCP analytes may potentiate the insulin amplification pathway via macronutrients or the incretin response.

## 4. Discussion

Our study shows that POPs are consistently detected in human pancreas, and for some pollutants, at higher concentrations than in adipose. We are the first to show that POP accumulation in human pancreas is associated with impaired beta cell function (data summarized in **Fig 8C**). Specifically, pancreas PCBs positively correlate with insulin secretion under low glucose conditions, whereas both PCBs and OCPs in pancreas negatively correlate with stimulation index (HG:LG ratio). Pancreas dioxins did not show any notable correlations with insulin secretion following a high glucose stimulus, but rather positively correlated with fatty acid- and amino acid-stimulated insulin secretion. These data imply that POP accumulation in pancreas can increase diabetes risk by impairing beta cell function, which could have implications on whole-body metabolic health. In fact, we also show that pancreas PCBs and OCPs positively correlate with systemic markers of diabetes risk, including age and BMI. Importantly, correlations between POPs and impaired beta cell function were not as prominent when using adipose analyte concentrations, emphasizing the importance of studying the pancreas in the context of metabolic disease. Overall, our data confirm that lipophilic pollutants accumulate in human pancreas tissues and positively correlate with markers of diabetes risk.

We report median dioxin/furan concentrations of 0.13 – 22.42 pg/g tissue, median PCB concentrations of 0.62 – 4,968 pg/g tissue, and median OCP concentrations of 10 – 35,200 pg/g tissue in adipose tissue from our study; these concentrations are towards the lower end of the concentration range reported in adipose samples from other countries. Specifically, median dioxin concentrations of 0.249 – 838 pg/g tissue were reported in adipose tissue from the USA general population in 1987 [73]; note that we could not find studies that assessed adipose dioxin concentrations (in wet weight) in the general population more recently than 1987. PCB levels of 41 – 4,186 pg/g tissue, 49,330 – 88,660 pg/g, and 10 – 4,310 pg/g tissue were reported in the USA (2007 – 2009) [74], Spain (2012 – 2014) [75] and China (2008 – 2009) [76], respectively. Lastly, adipose OCP concentrations in the range of 96 – 87,550 pg/g tissue, 119,400 – 303,830 pg/g tissue, and 30 – 3,598,000 pg/g were reported in the USA (2012 – 2014) [75], Spain (2008 – 2009) [76] and China (2008 – 2009) [76], respectively. Our data suggest that POP levels in human tissues are modestly decreasing over time, potentially due to the implementation of regulatory actions to reduce the use and industrial release of POPs. Regardless, we are still detecting high levels of POPs in human tissues, which could have detrimental effects on health.

The most concentrated OCPs in adipose and pancreas reported in our study were 4,4’-DDE, hexachlorobenzene, trans-nonachlor, oxychlordane, dieldrin, and β-HCH. Our data are in line with human biomonitoring studies in Canada [77] and the USA [78], which reported that 4,4’-DDE, hexachlorobenzene, β-HCH, trans-nonachlor, and oxychlordane are the most abundant OCP analytes in human serum; 4’-DDE concentrations were reported at levels ∼20x higher than any other OCP analyte [77,78]. 4,4’-DDE is the metabolite of 4,4’-DDT. Although DDT is banned in Canada [77], it is still used in developing countries for disease control [40]; thus DDT is being released into our environment, and given its persistent nature, the long-range atmospheric transport of this compound can lead to global contamination in our water and soil [79–81]. Our data indicate that Canadians are still exposed to DDT, which could have important health implications. The most abundant dioxins/furans in human pancreas and adipose from our study were OCDD, 1,2,3,4,6,7,8-HPCDD, and 1,2,3,4,6,7,8-HXCDD, and the most abundant PCBs were PCB 180+193, PCB 118, PCB 170, and PCB 156+157. These findings are also in line with biomonitoring studies in Canada and the USA [4,77,82]. Very little is known about the metabolic health risks associated with these specific POP analytes. Importantly, we provide novel evidence that dioxins/furans, PCBs and OCP levels in human pancreas correlate with different markers of diabetes risk.

Our perifusion data revealed that pancreatic concentrations of PCBs and OCPs were generally correlated with measures of beta cell dysfunction (data summarized in **Fig 8C**). Pancreas PCBs and OCPs showed weak to moderate positive correlations with basal insulin secretion under LG conditions. Basal hypersecretion of insulin can be an early indicator of metabolic dysfunction; chronic overactivation of beta cells can lead to beta cell exhaustion and apoptosis, leading to T2D [83,84]. Hyperinsulinemia can also drive the development of obesity [85]; this aligns with the positive correlations observed between pancreas PCBs and OCPs and BMI. We also present compelling evidence that pancreas PCBs and OCPs negatively correlate with stimulation index, irrespective of the research lab, analysis technique, and glucose concentrations used in the analysis, further suggesting that PCBs and OCPs impair beta cell function. Given that beta cell dysfunction is a hallmark of T2D pathogenesis [83], our data points to PCB/OCP exposure as a risk factor for developing T2D. These data support epidemiological studies showing a negative association between serum PCBs [42,44,86] and OCPs [42,44,86] with glucose-stimulated plasma insulin and HOMA-β (homeostasis model assessment of beta cell function). We also identified a subset of OCP analytes in adipose and pancreas that positively correlated with %HbA1c, further supporting an association between pancreas OCP accumulation and increased T2D risk. However, it is important to note that only a small subset of the donors used in this analysis had %HbA1c levels outside the normal range (i.e. %HbA1c > 5.7; n = 6), thus limiting our ability to elucidate correlations between POP concentrations and blood glucose levels. Studies with a larger sample size that includes both non-T2D and T2D donors are warranted to better understand this relationship.

We found that POP concentrations are generally ∼63% lower in pancreas than adipose, except for 1,2,3,4,6,7,8-HPCDD, 1,2,3,4,6,7,8-HPCDF, 1,2,3,7,8,9-HXCDF, OCDF, and PCB 77. These data suggest that certain POP analytes may be preferentially sequestered into the pancreas and/or are excreted from the pancreas more slowly, which may be of particular concern for pancreas/islet health and consequently metabolic disease. Surprisingly, we did not observe any correlations between 1,2,3,4,6,7,8-HPCDD, 1,2,3,4,6,7,8-HPCDF, 1,2,3,7,8,9-HXCDF, and OCDF with donor characteristics, basal insulin secretion or GSIS. However, 1,2,3,4,6,7,8-HPCDF and OCDF are amongst the dioxin/furan analytes showing the strongest positive correlations with fatty acid- and amino acid-stimulated insulin secretion. In addition, correlations with PCB 77 differed from all other PCBs; PCB 77 did not show any significant correlations with donor characteristics or GSIS, but showed a moderate positive correlation with fatty acid- and amino acid-stimulated insulin secretion. These findings are particularly interesting since fatty acids [87,88] and branched-chain amino acids (such as leucine) [89–91] are known to potentiate insulin secretion, suggesting that a subset of POPs—mainly analytes that preferentially accumulate in pancreas compared to adipose—cause islet dysfunction by altering insulin amplification pathways. This highlights the need to consider individual analytes when assessing the effects of POPs on metabolic health and beta cell function, rather than relying solely on pollutant class-level summary measures.

It is important to note both strengths and limitations to our study. An important strength of this study is the unique access to isolated primary human islets and tissue biopsies from the same donors. This allowed for direct and detailed measurements of nutrient-stimulated insulin secretion, as opposed to indirect measures in plasma samples (like HOMA-β or fasting plasma insulin). The reported correlations between pancreas PCBs/OCPs with beta cell dysfunction are particularly compelling given that the correlations were maintained when beta cell function was measured by different techniques in different research labs. Our sample size of 31 donors is the largest report of POP measurements in human pancreas to-date; however, this sample size is relatively small for the purposes of capturing heterogeneity in POP levels and beta cell function among donors. Specifically, our small size prevented us from stratifying our data based on diabetes status to assess correlations between POP concentrations and beta cell function in non-T2D vs T2D donors. Additionally, there is evidence that POPs increase diabetes risk in a sex-specific manner [92]. As such, we visualized the spread of our female and male donors for key correlations using scatter plots, but our sample sizes were not sufficient to statistically test the effect of sex in our cohort. Future studies should build on our findings to assess whether similar correlations are found with a bigger cohort of donors and whether any of these correlations are dependent on diabetes status or sex.

We focused on 3 classes of POPs that are largely regulated in Canada and the USA. It would be valuable to measure POPs that are not well regulated, such as per- and polyfluoroalkyl substances (PFAS), which were recently added to the Stockholm Convention [93]. PFAS are widely detected in serum [94,95] and maternal milk [96], and have been associated with a variety of health complications [97–99]. However, the extent of accumulation in human tissues, and whether PFAS concentrations are correlated with beta cell function is unclear. Lastly, our POP analysis is presented as wet weight concentrations rather than lipid-normalized due to insufficient tissue material to measure lipid levels. Given that POPs are highly lipophilic, it will be important to assess whether POP concentrations and correlations with beta cell function parameters are influenced by lipid content in the tissue. Regardless of these limitations, our study shows convincing evidence that dioxins/furans, PCBs, and OCPs are detected in human adipose and pancreas, although concentrations were generally lower than previously reported in other studies. Importantly, we are the first to show that dioxin/furan, PCB and OCP concentrations in pancreas are consistently correlated with measures of beta cell dysfunction in isolated human islets.

## Data Availability

All data produced in the present study are available upon reasonable request to the authors.

## Acknowledgements

This research was supported by a Canadian Institutes of Health Research (CIHR) Project Grant (#PJT-2018-159590). J.E.B. is supported by an Early Researcher Award from the Ontario Government. E.E.M is supported by a Diabetes Canada End Diabetes Award (OG-3-24-5818-EM). M.P.H. was supported by a CIHR CGS-D award. M.E.A.C. was supported by an NSERC CGS-M and NSERC CGS-D award. J.P. was supported by the Guiding interdisciplinary Research on Women’s and girls’ health and Wellbeing (GROWW) scholarship and Ontario Graduate Scholarship.

This work includes data from HumanIslets.com funded by the Canadian Institutes of Health Research, JDRF Canada, and Diabetes Canada (5-SRA-2021-1149-S-B/TG 179092) with data from islets isolated by the Alberta Diabetes Institute IsletCore with the support of the Human Organ Procurement and Exchange program, Trillium Gift of Life Network, BC Transplant, Quebec Transplant, and other Canadian organ procurement organizations.

## 5. Author Contributions

M.P.H. and J.E.B. conceived the experimental design and wrote the manuscript. M.P.H., M.E.A.C., J.P., E.F., J.K., J.L., N.S., E.E.M., F.C.L., J.D.J., J.E.M.F., P.E.M., and J.E.B. were involved with data acquisition, data analysis, and interpretation of data; J.A-M., was involved in data analysis and interpretation. All authors contributed to manuscript revisions and approved the final version of the article.

## Abbreviations

AhR: Aryl hydrocarbon receptor
CYP1A1: Cytochrome P450 1A1
DL: Dioxin-like
NDL: Non-dioxin-like
OCPs: Organochlorine pesticides
PCBs: Polychlorinated biphenyls
PCDDs: Polychlorodibenzo-*p*-dioxins
PCDFs: Polychlorodibenzofurans
POPs: Persistent organic pollutants
T2D: Type 2 diabetes
TCDD: 2,3,7,8-tetrachlorodibenzo-*p*-dioxin

