## Supplementary Material for "Persistent organic pollutant concentrations in human pancreas and peripancreatic adipose tissues correlate with markers of beta cell dysfunction"

**SUPPLEMENTAL TABLES**

**Supp. Table 1**: **Donor characteristics for all human organ donors used in this analysis**. Age range, BMI, % HbA1c, diagnosis, and diabetes status. Diabetes status is relative to %Hba1c rather than clinical diagnosis. F = female, M = male, T2D = type 2 diabetes. All donor characteristics are publicly available via the IsletCore website ([www.humanislets.com](http://www.humanislets.com)).

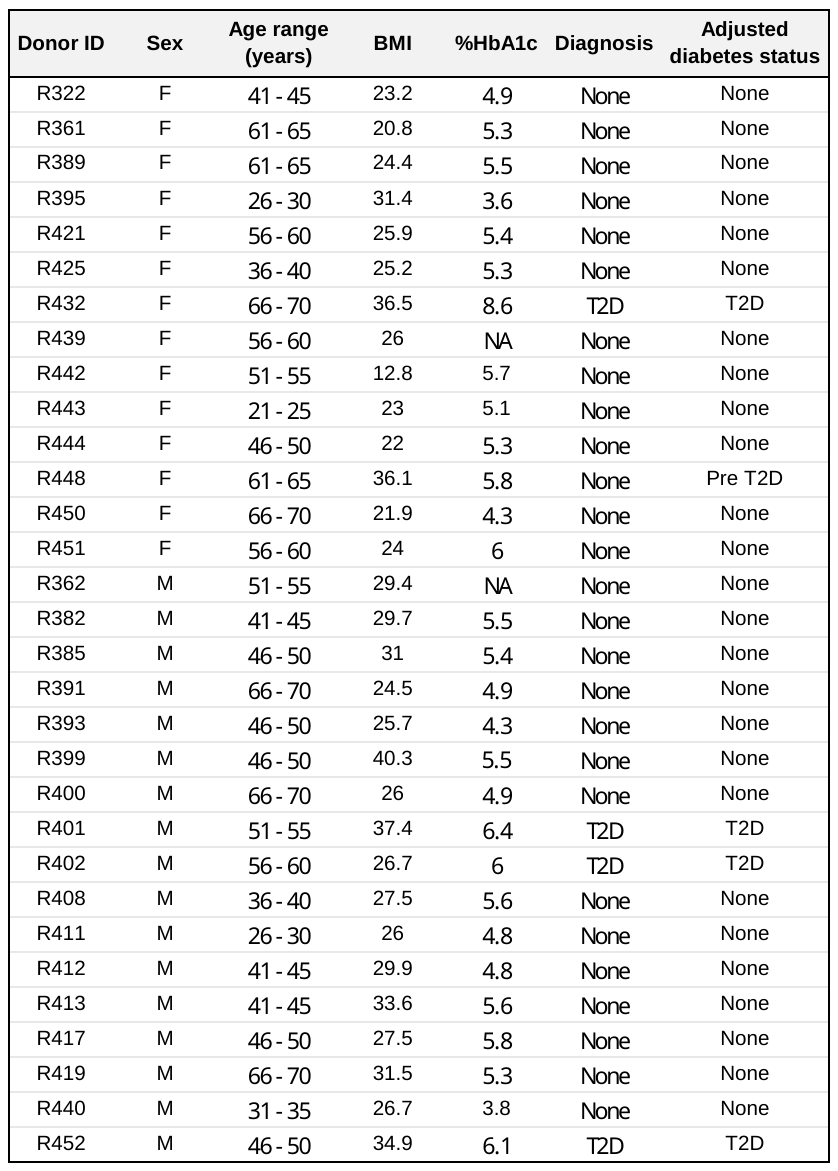

**Supp. Table 2**: Summary of individual donor functional assessments retained for correlation analysis. Functional assessments were conducted in 3 individual labs, including perifusion in the Bruin lab and Johnson lab, and static glucose-stimulated insulin secretion analysis by the IsletCore. LG = low glucose, HG = high glucose, Ex4 = exendin4, AUC = area under the curve.

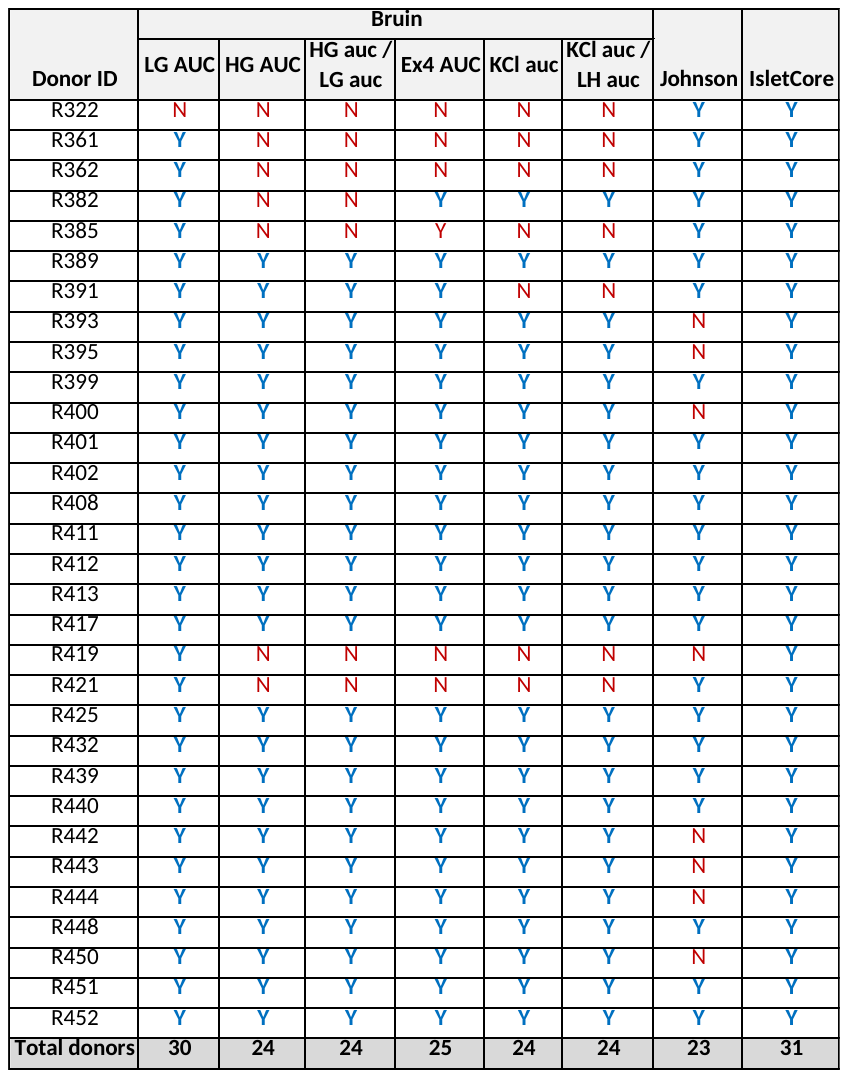

**SUPPLEMENTAL FIGURES**

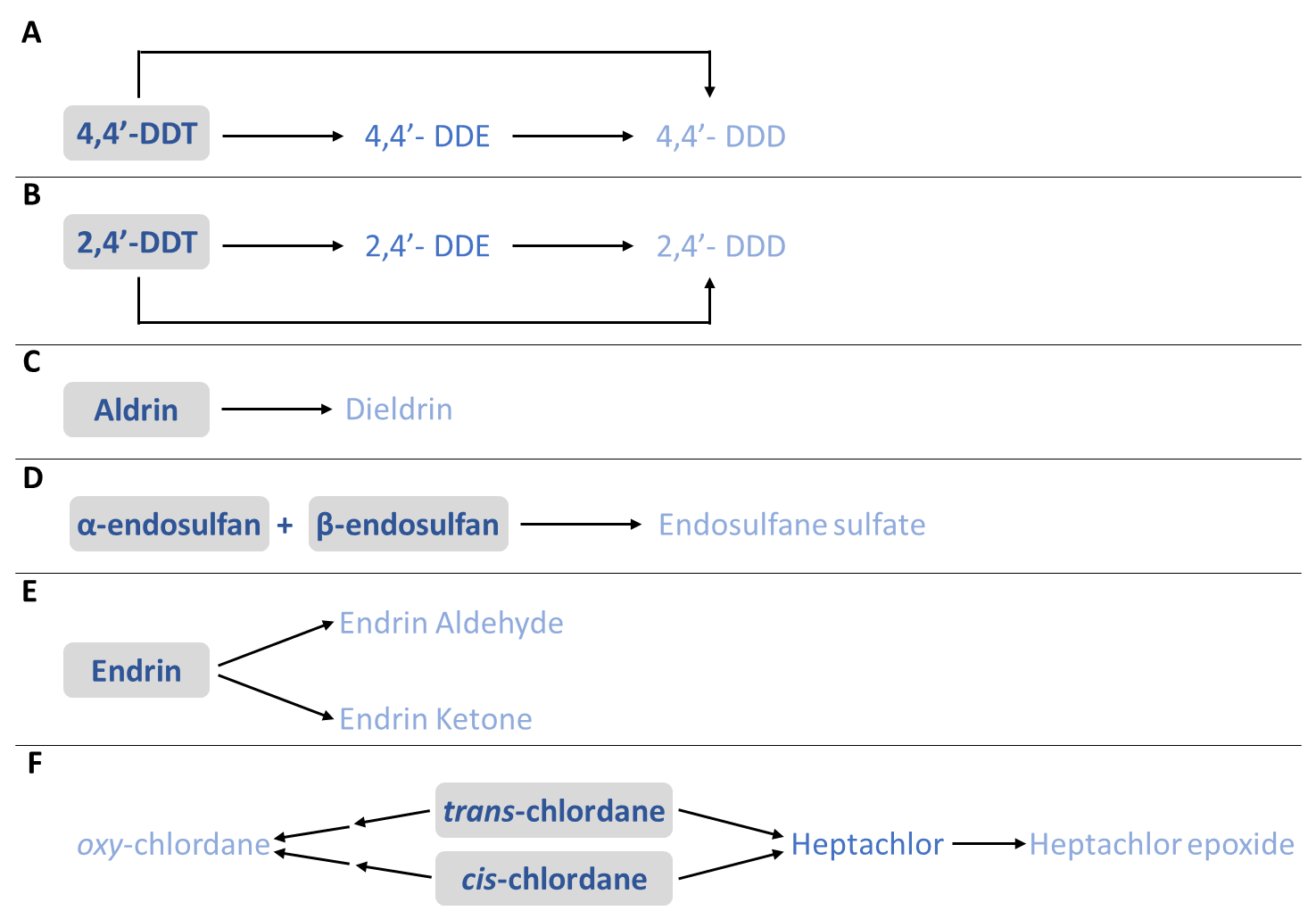

**Supp. Fig. 1: Overview of organochlorine pesticides analysed and their degradation pathways. (A)** 4,4’-DTT is converted to 4,4’-DDE and 4,4’-DDD. **(B)** 2,4’-DTT is converted to 2,4’-DDE and 2,4’-DDD. **(C)** Aldrin is converted to dieldrin. **(D)** α-endosulfan and β-endosulfan react to form endosulfane sulfate. **(E)** Endrin is converted to endrin aldehyde and/or endrin ketone. **(F)** *trans*-chlordane and *cis*-chlordane are converted to oxy-chlordane or heptachlor; heptachlor is further converted to heptachlor epoxide.

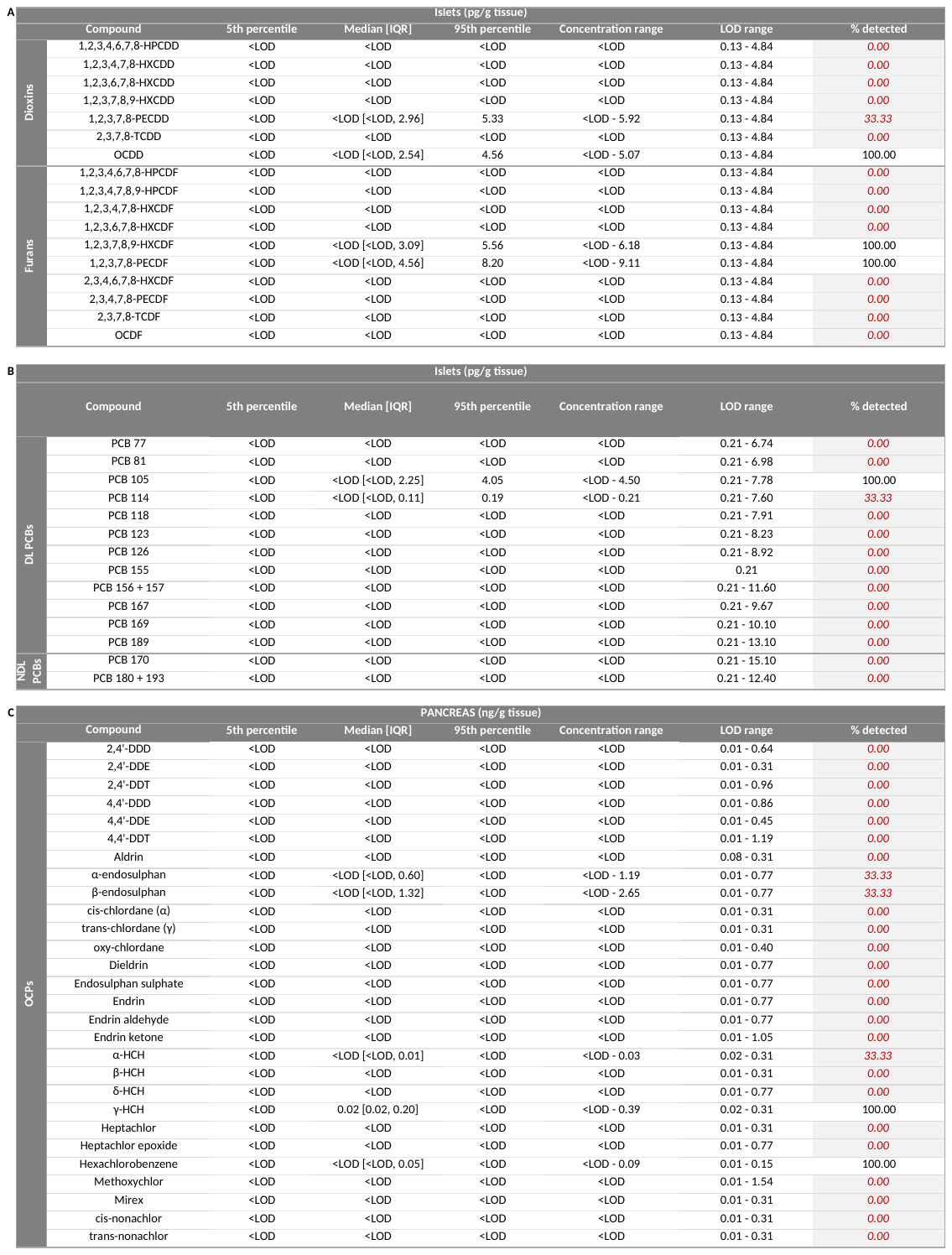

**Supp. Fig. 2: Descriptive statistics of dioxins/furans, polychlorinated biphenyls (PCBs), and organochlorine pesticides (OCPs) in human islets.** Dioxins/furans, PCBs and OCPs were measured in 4000 human islets from 3 human organ donors. Descriptive statistics for all **(A)** dioxin/furan (pg/g tissue), **(B)** dioxin-like (DL) and non-dioxin-like (NDL) PCBs (pg/g tissue), and **(C)** OCP (ng/g tissue) analytes measured in islets. IQR = interquartile range; min = minimum concentration found; max = maximum concentration found; LOD = sample limit of detection. IQR range represents the 25^th^ to 75^th^ percentile. All data represents raw un-corrected concentrations. Analytes that were detected in <50% of donors are indicated by red text with grey highlighting.

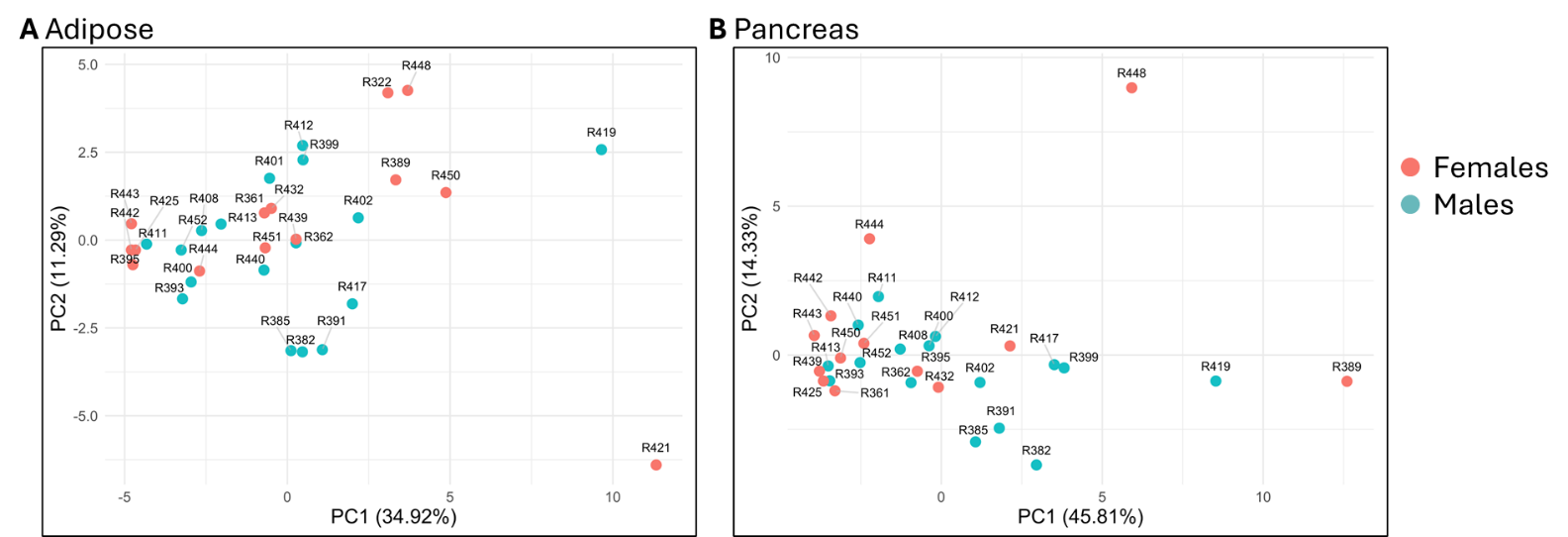

**Supp. Fig. 3: Pollutant concentration profiles differed for 2 donors in adipose and 3 donors in pancreas.** Donors with differing tissue-level pollutant concentration profiles were identified using principal components analysis (PCA). PCA was performed based on dioxin/furan, PCB, and OCP concentrations in **(A)** adipose or **(B)** pancreas tissues. Each point represents a donor and is labelled by donor ID. The analyses were performed using the pcaMethods package (version 1.98.0) with the NIPALS algorithm for missing value imputation.

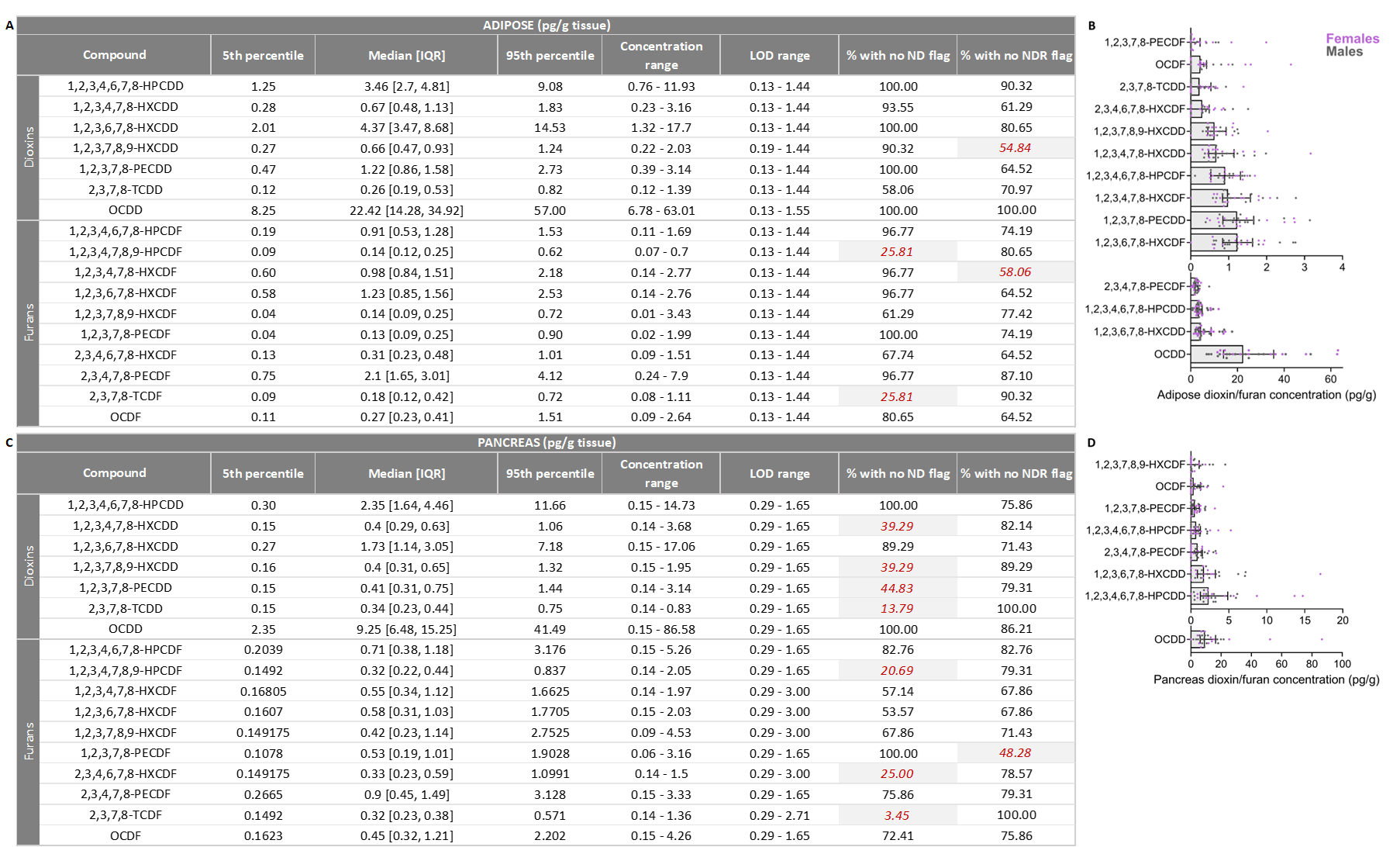

**Supp. Fig. 4: Descriptive statistics of dioxins/furans in human adipose and pancreas tissue.** All concentrations are reported as pg/g tissue. Descriptive statistics for all dioxin/furan analytes measured in **(A)** adipose and **(C)** pancreas. IQR = interquartile range; min = minimum concentration found; max = maximum concentration found; LOD = sample limit of detection; ND = not detected; NDR = peak detected but did not meet quantification criteria. IQR range represents the 25^th^ to 75^th^ percentile. Of the 17 dioxin/furan analytes measured, **(A)** 4 analytes in adipose and **(C)** 8 analytes in pancreas did not meet the inclusion criteria and were removed from the analysis (indicated by the red text with grey highlighting). **(B,D)** Dioxin/furan concentrations for analytes that met all inclusion criteria and were retained for analysis. Data points represent individual human organ donors, colour coded by sex. All data are presented as median +/- interquartile range. All data presented were blank corrected and any zeros were assigned a value of 1/2 LOD.

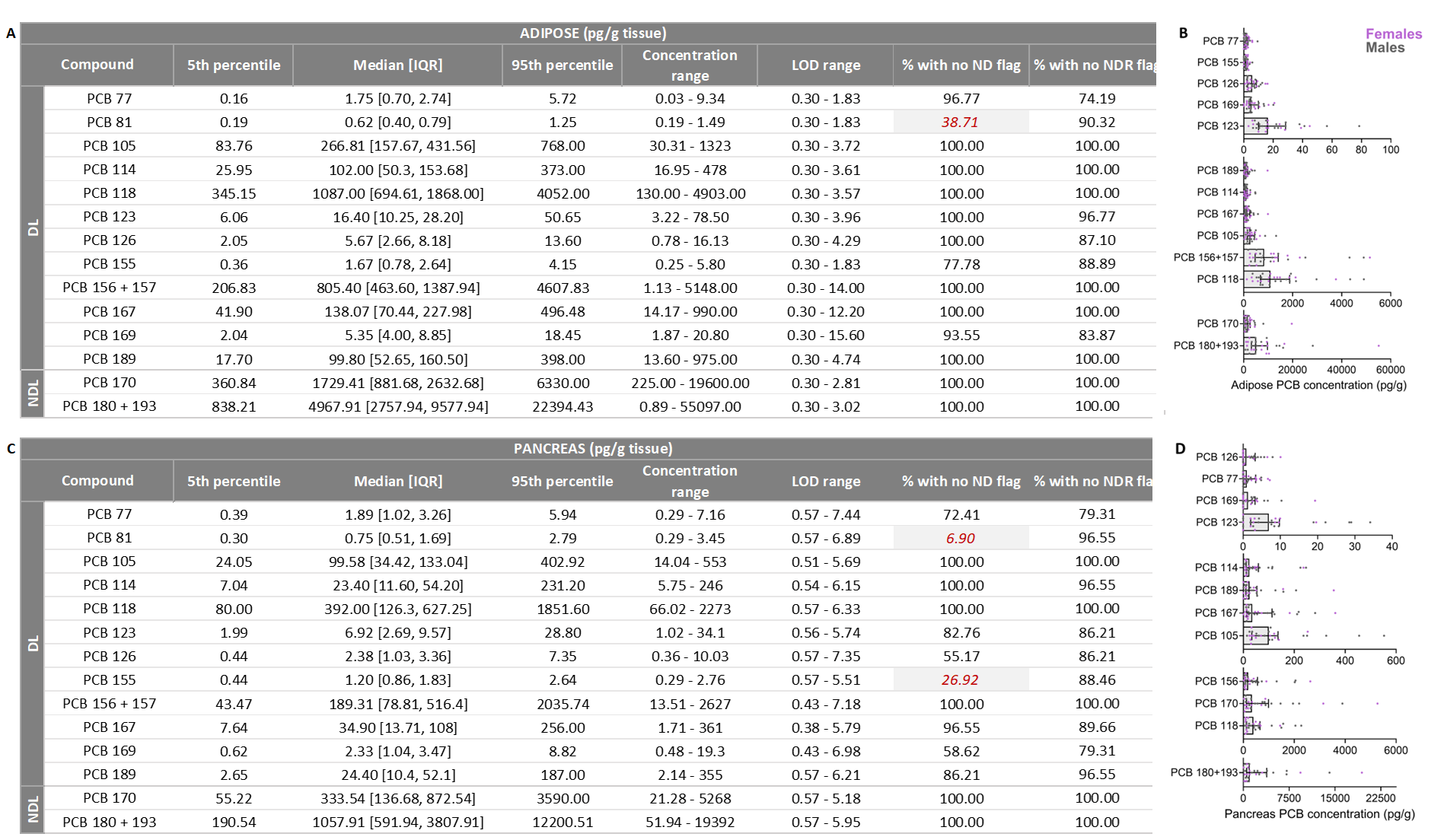

**Supp. Fig. 5: Descriptive statistics of polychlorinated biphenyls (PCBs) in human adipose and pancreas tissue.** All concentrations are reported as pg/g tissue. Descriptive statistics for all PCB analytes measured in **(A)** adipose and **(C)** pancreas, including dioxin-like (DL) and non-dioxin-like (NDL) PCBs. IQR = interquartile range; min = minimum concentration found; max = maximum concentration found; LOD = sample limit of detection; ND = not detected; NDR = peak detected but did not meet quantification criteria. IQR range represents the 25^th^ to 75^th^ percentile. Of the 16 PCB analytes measured, **(A)** 1 analyte in adipose and **(C)** 2 analytes in pancreas did not meet the inclusion criteria and were removed from the analysis (indicated by the red text with grey highlighting). **(B,D)** PCB concentrations for analytes that met all inclusion criteria and were retained for analysis. Data points represent individual human organ donors, colour coded by sex. All data are presented as median +/- interquartile range. All data presented were blank corrected and any zeros were assigned a value of 1/2 LOD.

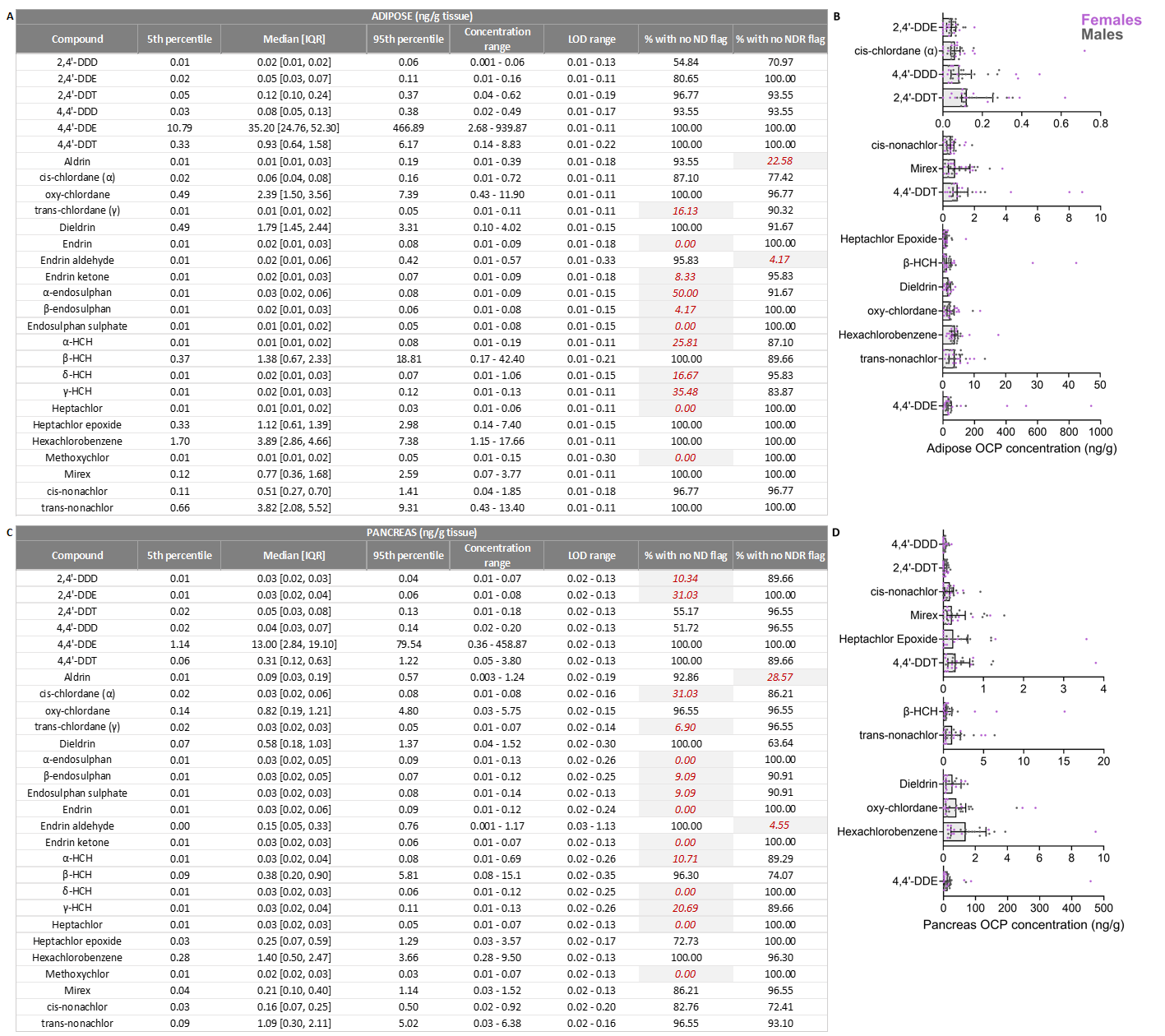

**Supp. Fig. 6: Descriptive statistics of organochlorine pesticides (OCPs) in human adipose and pancreas tissue.** All concentrations are reported as ng/g tissue. Descriptive statistics for all OCP analytes measured in **(A)** adipose and **(C)** pancreas. IQR = interquartile range; min = minimum concentration found; max = maximum concentration found; LOD = sample limit of detection; ND = not detected; NDR = peak detected but did not meet quantification criteria. IQR range represents the 25^th^ to 75^th^ percentile. Of the 28 OCP analytes measured, **(A)** 13 analyte in adipose and **(C)** 16 analytes in pancreas did not meet the inclusion criteria and were removed from the analysis (indicated by the red text with grey highlighting). **(B,D)** OCP concentrations for analytes that met all inclusion criteria and were retained for analysis. Data points represent individual human organ donors, colour coded by sex. All data are presented as median +/- interquartile range. All data presented were blank corrected and any zeros were assigned a value of 1/2 LOD.

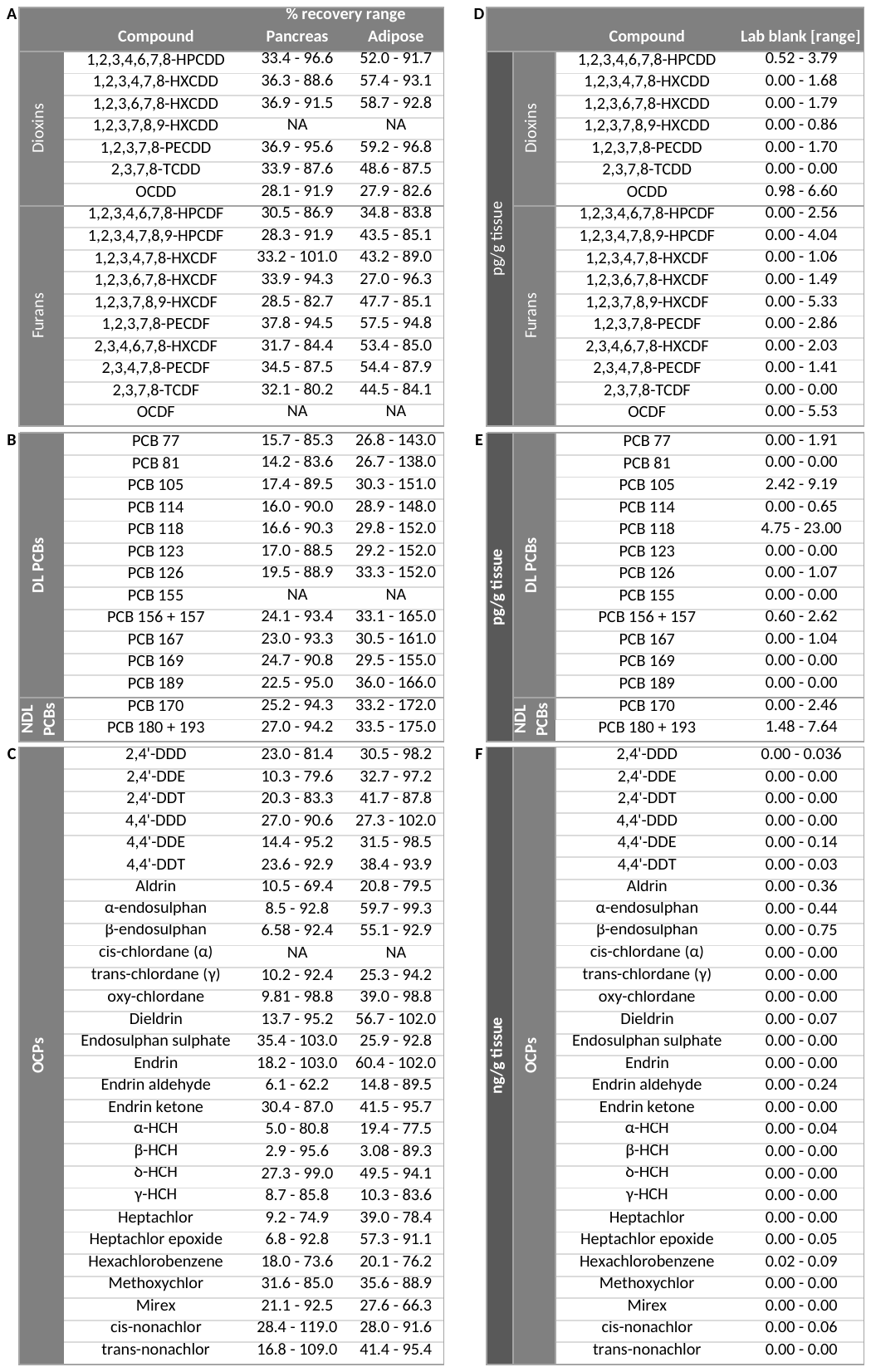

**Supp. Fig. 7: Percent recovery and lab blank concentration ranges for dioxin/furan, polychlorinated biphenyl (PCB), and organochlorine pesticide (OCP) analytes. (A-C)** Percent recoveries for **(A)** dioxins/furans, **(B)** dioxin-like (DL) and non-dioxin-like (NDL) PCBs, and **(C)** OCPs. All samples were spiked with ^13^C-labelled surrogate standards and % recovery was calculated for each surrogate. **(D-F)** Concentrations of **(D)** dioxins/furans, **(E)** PCBs, and **(F)** OCPs in lab blanks. Dioxin/furan and PCB concentrations are presented as pg/g tissue, OCPs are presented as ng/g tissue.

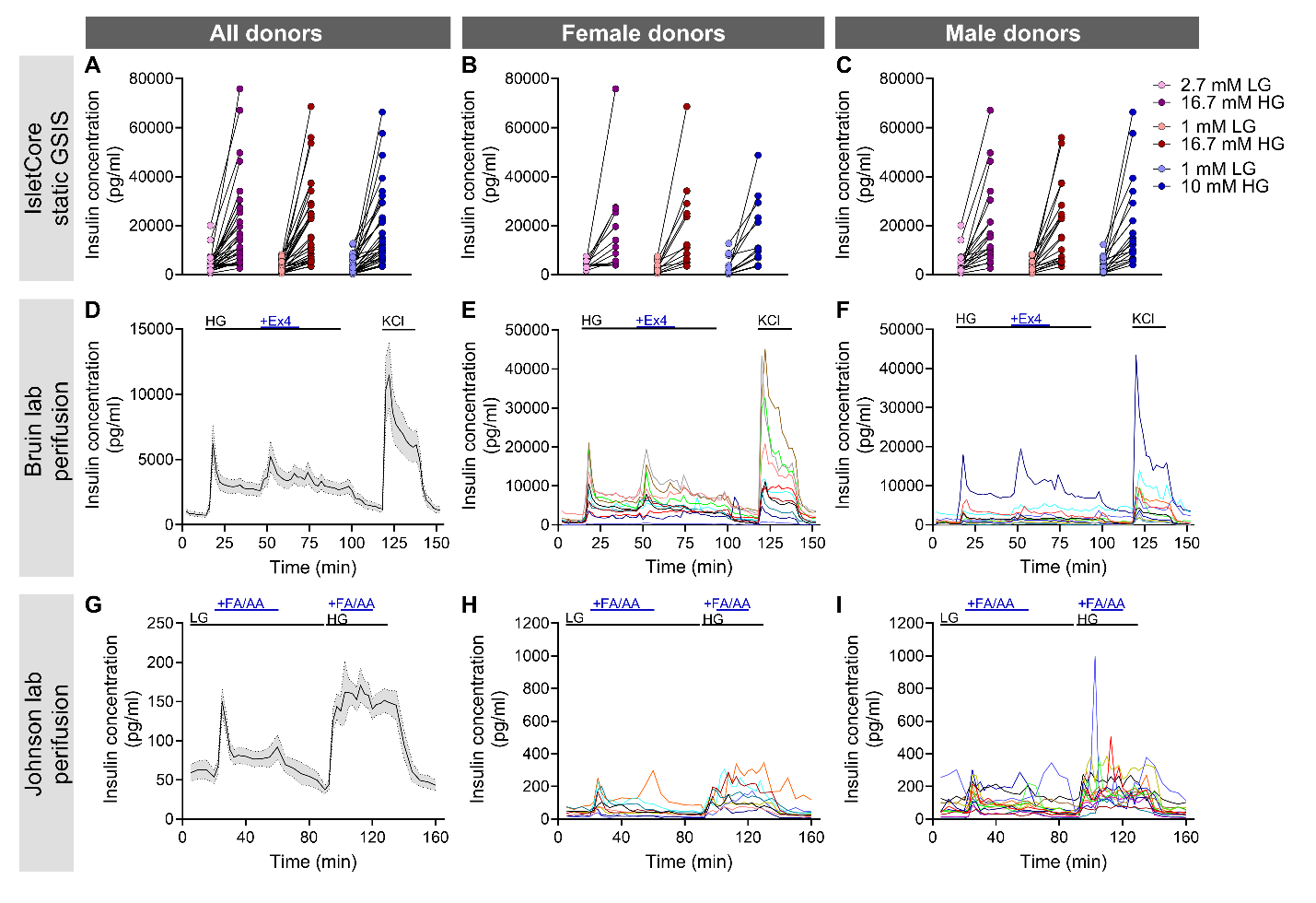

**Supp. Fig. 8**: **Nutrient-stimulated insulin secretion from human organ donor islets**. Function assessments were conducted **(A-C)** at the Alberta Diabetes Institute IsletCore, **(D-F)** in the lab of Dr. Jennifer Bruin, and **(G-I)** in the lab of Dr. James Johnson using **(A-C)** static glucose-stimulated insulin secretion (GSIS) or **(D-I)** dynamic perifusion analysis. LG = low glucose, HG = high glucose, Ex4 = exendin 4, FA = fatty acid, AA = amino acid. We assessed islet function in a total of 31 human donors, of which 22 donors were assessed by all three labs, 8 donors were assessed by the Bruin lab and IsletCore only, and 1 donor was assessed by the Johnson lab and IsletCore only. **(A-C)** Static insulin secretion in response to sequential incubations with 2.7 mM LG and 16.7 mM HG, 1 mM LG and 16.7 mM HG, or 1 mM LG and 10 mM HG; each GSIS condition was conducted on separate subsets of islets. **(D-F)** Dynamic insulin secretion in response to 2.8 mM LG, 10 mM HG, 10mM HG + 100nM Ex4, and 30 mM KCl. **(G-I)** Dynamic insulin secretion in response to 3 mM LG, 3 mM + 5mM leucine or 1.5mM oleate/palmitate (1:1 mix), 6 mM HG, and 6 mM HG + 5mM leucine or 1.5mM oleate/palmitate (1:1 mix). **(A-C)** Data are presented as before-after plots for **(A)** all donors, **(B)** females only, and **(C)** males only; data points connected by a line represent individual donors. **(D-I)** Data are presented as **(D,G)** mean +/- SEM of all donor data, and **(E,F,H,I)** individual donor perifusion curves, separated by **(E,H)** female and **(F,I)** male data.

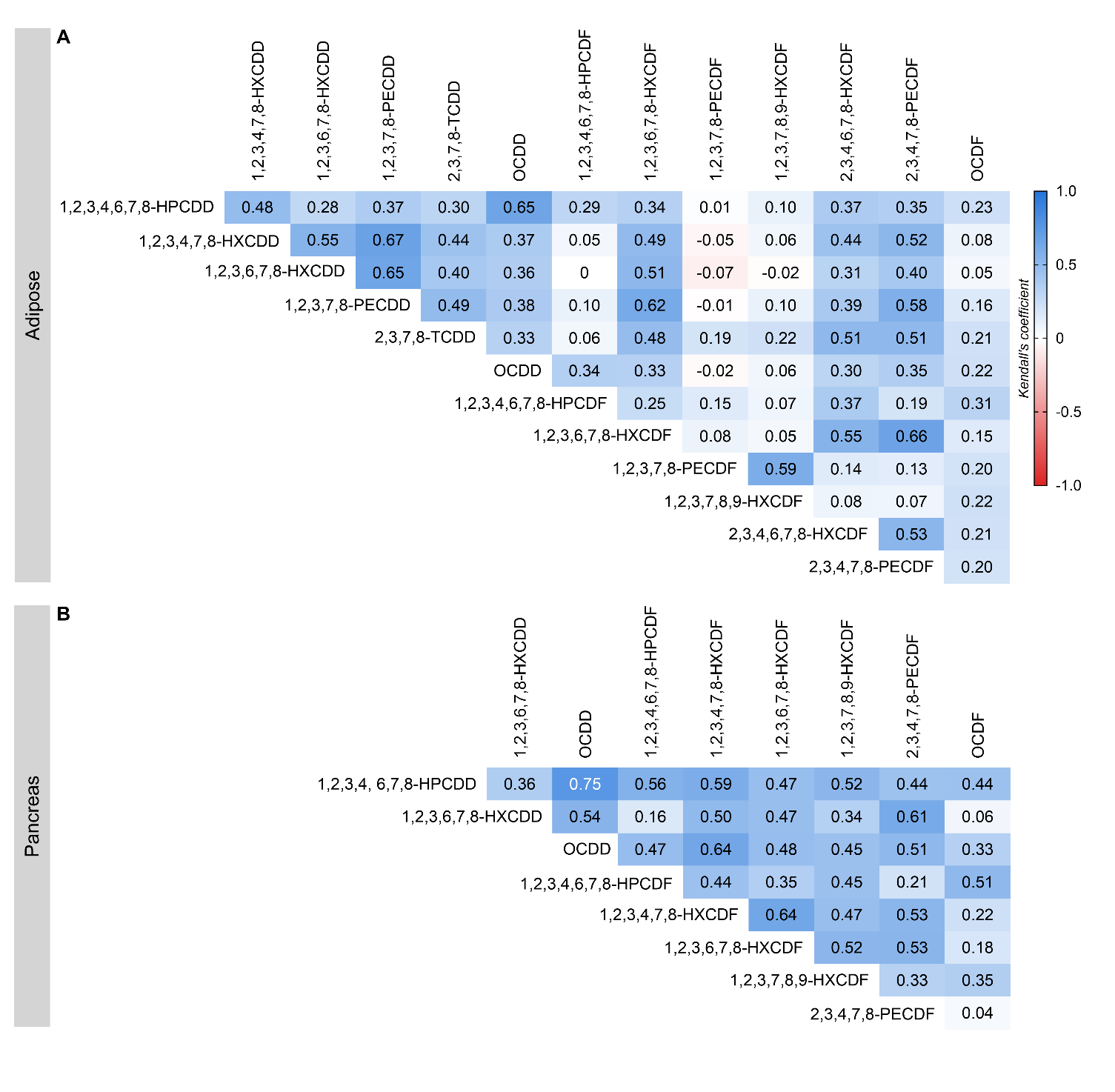

**Supp. Fig. 9**: **Dioxin/furan analytes largely show positive correlations with each other within a tissue**. Heatmaps depicting correlations between dioxin/furan analytes within **(A)** adipose and **(B)** pancreas. All analyte concentrations used in this analysis were blank corrected and any zeros were assigned a value of 1/2 LOD. Data in the heatmaps represent Kendall’s rank correlation coefficients.

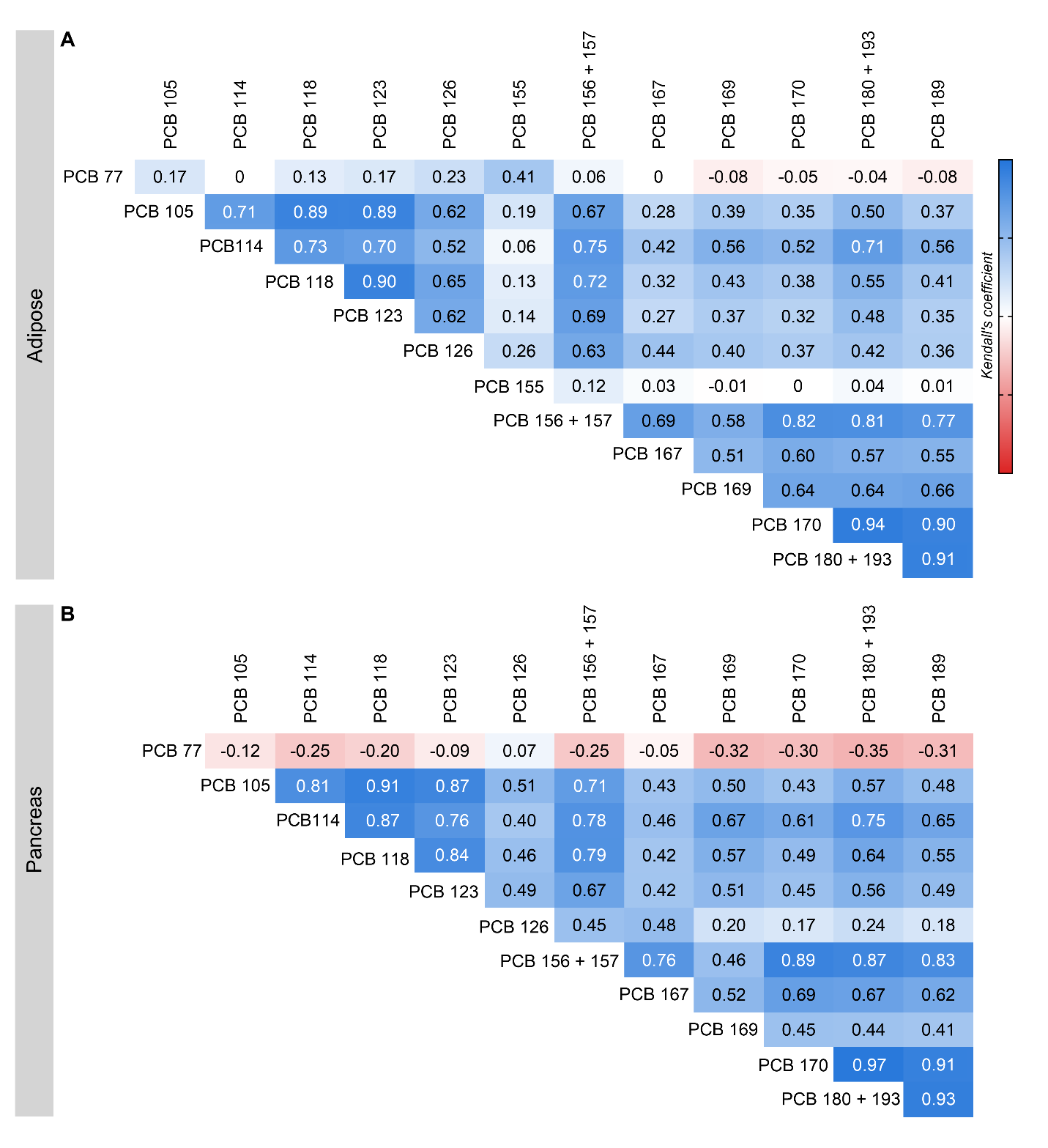

**Supp. Fig. 10**: **PCB analytes largely show positive correlations with each other within a tissue, with the exception of PCB 77**. Heatmaps depicting correlations between polychlorinated biphenyls (PCBs) analytes within **(A)** adipose and **(B)** pancreas. All analyte concentrations used in this analysis were blank corrected and any zeros were assigned a value of 1/2 LOD. Data in the heatmaps represent Kendall’s rank correlation coefficients.

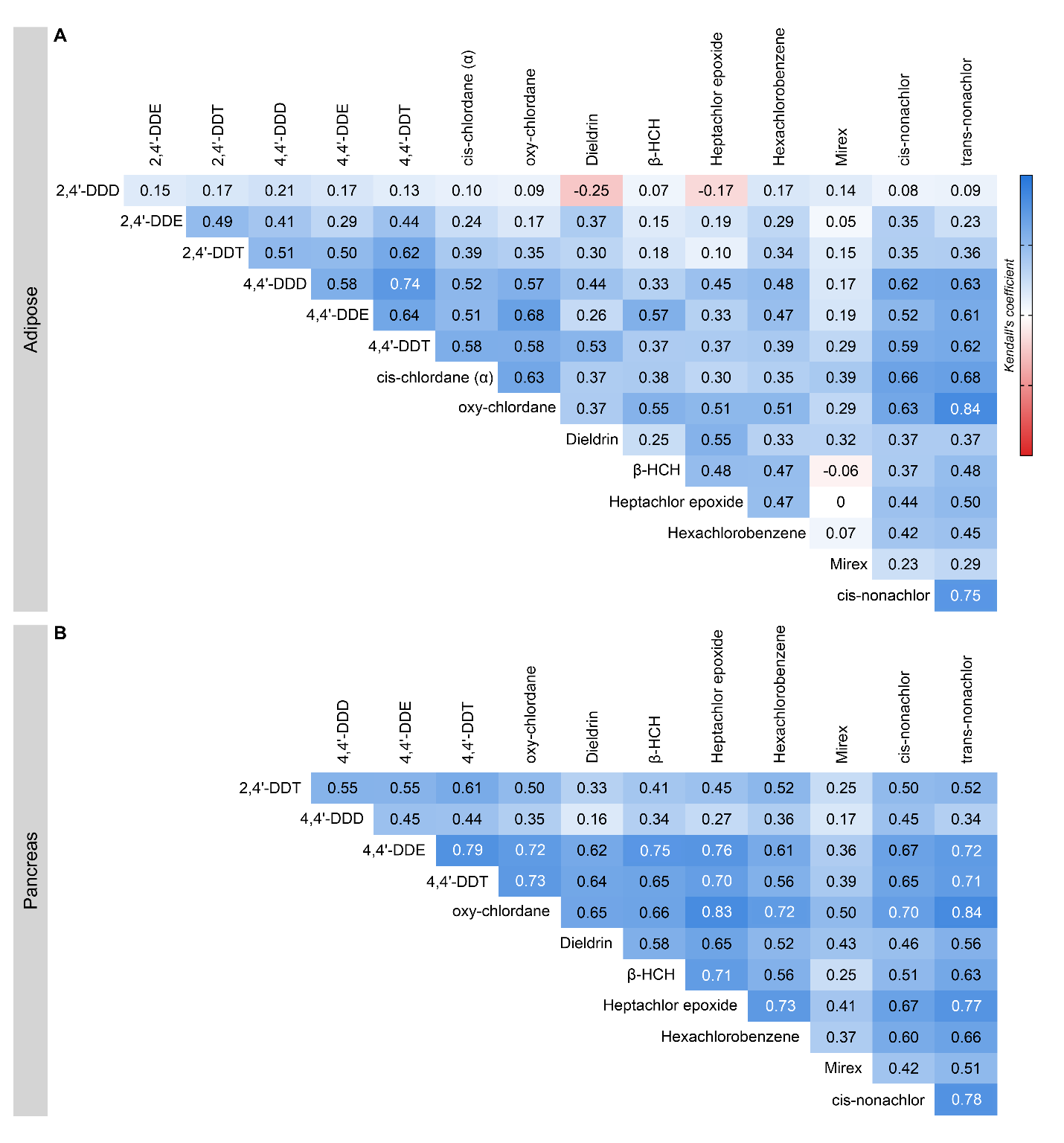

**Supp. Fig. 11**: **OCP analytes largely show positive correlations with each other within a tissue**. Heatmaps depicting correlations between organochlorine pesticide (OCP) analytes within **(A)** adipose and **(B)** pancreas. All analyte concentrations used in this analysis were blank corrected and any zeros were assigned a value of 1/2 LOD. Data in the heatmaps represent Kendall’s rank correlation coefficients.

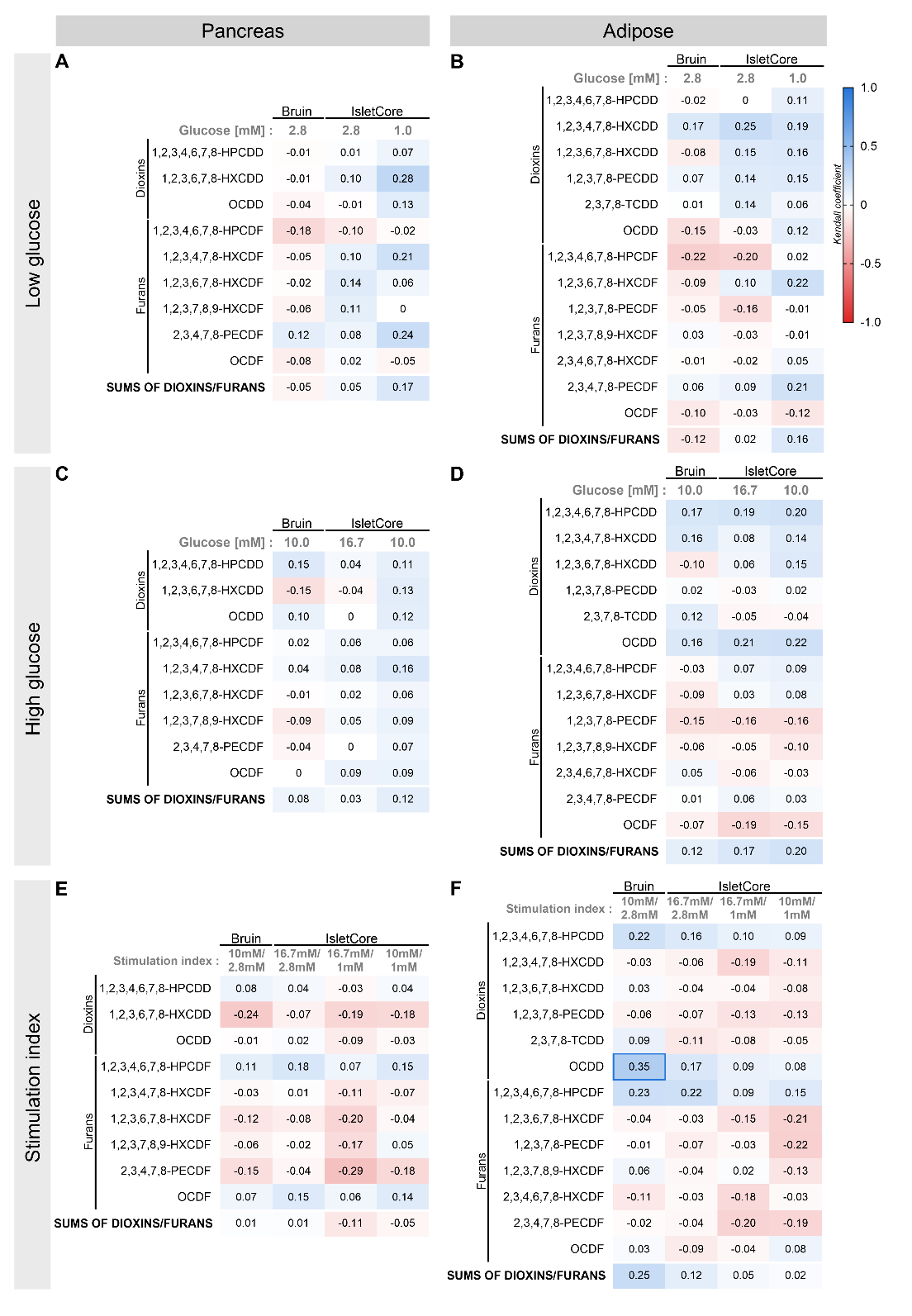

**Supp. Fig. 12**: **Dioxin/furan concentrations in human pancreas and adipose did not correlate with insulin secretion under low- or high-glucose conditions.** Heatmaps depicting correlations between dioxin/furan concentrations in human donor **(A,C,E)** pancreas and **(B,D,F)** adipose tissue and islet function. Human donor islets were stimulated with **(A,B)** 1.0 or 2.8 mM low glucose (LG), and **(C,D)** 10.0 or 16.7 mM high glucose (HG), and total insulin secretion was measured following each stimulus. **(E,F)** Stimulation index was calculated as a ratio of insulin concentration under HG relative to LG condition. All analyte concentrations used in this analysis were blank corrected and any zeros were assigned a value of 1/2 LOD. Data in the heatmaps represent Kendall’s rank correlation coefficients. Bolded boxes emphasize correlations that are moderate to strong.

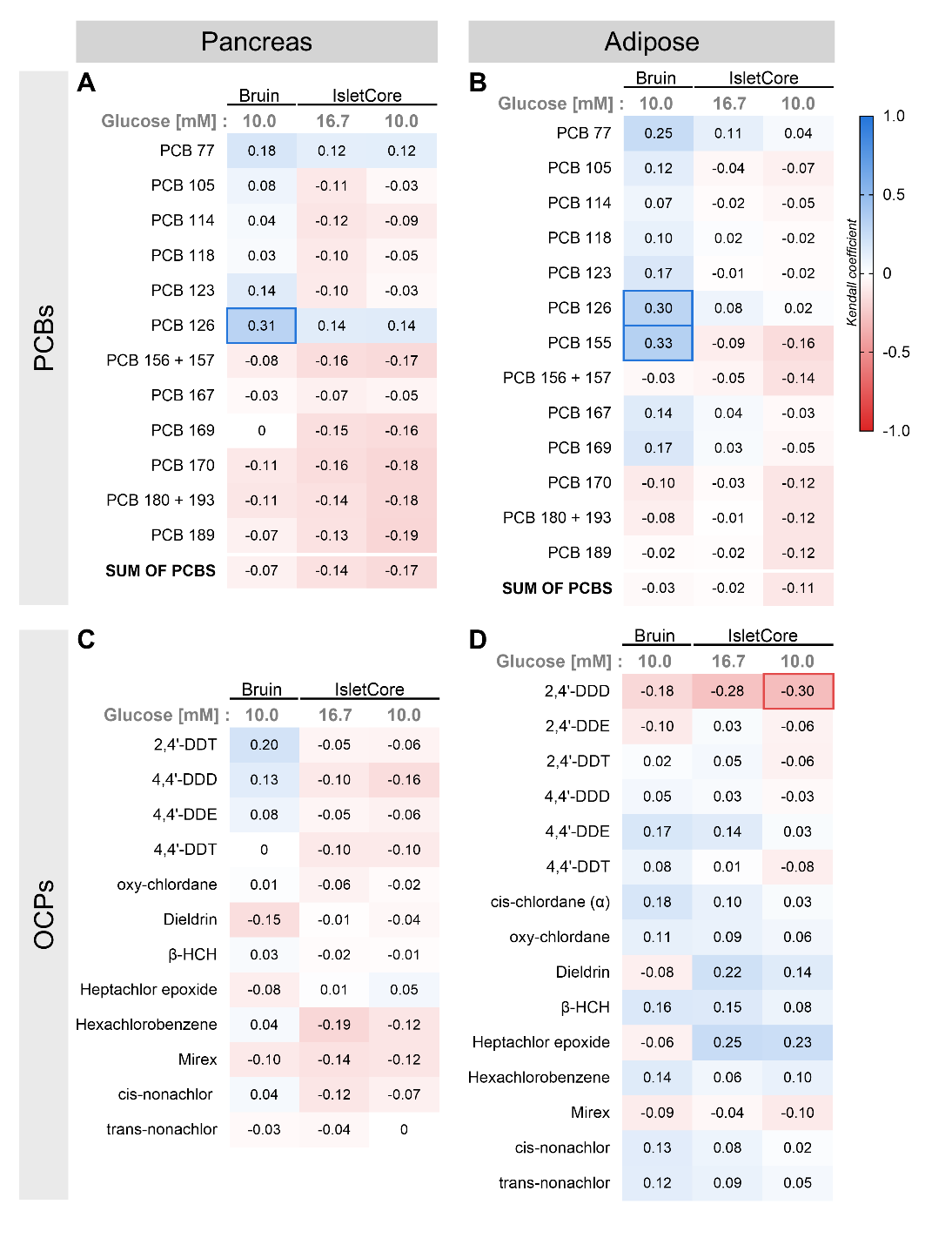

**Supp. Fig. 13: PCB and OCP concentrations in pancreas and adipose showed no notable correlations with insulin secretion under high glucose conditions.** Heatmaps depicting correlations between **(A,B)** polychlorinated biphenyls (PCBs) and **(C,D)** organochlorine pesticides (OCPs) concentrations in human donor **(A,C)** pancreas and **(B,D)** adipose and insulin secretion under high glucose (HG) conditions. Human donor islets were stimulated with 10 mM or 16.7 mM glucose. Total insulin secretion was measured following each stimulus. All analyte concentrations used in this analysis were blank corrected and any zeros were assigned a value of 1/2 LOD. Data in the heatmaps represent Kendall’s rank correlation coefficients. Bolded boxes emphasize correlations that are moderate to strong.

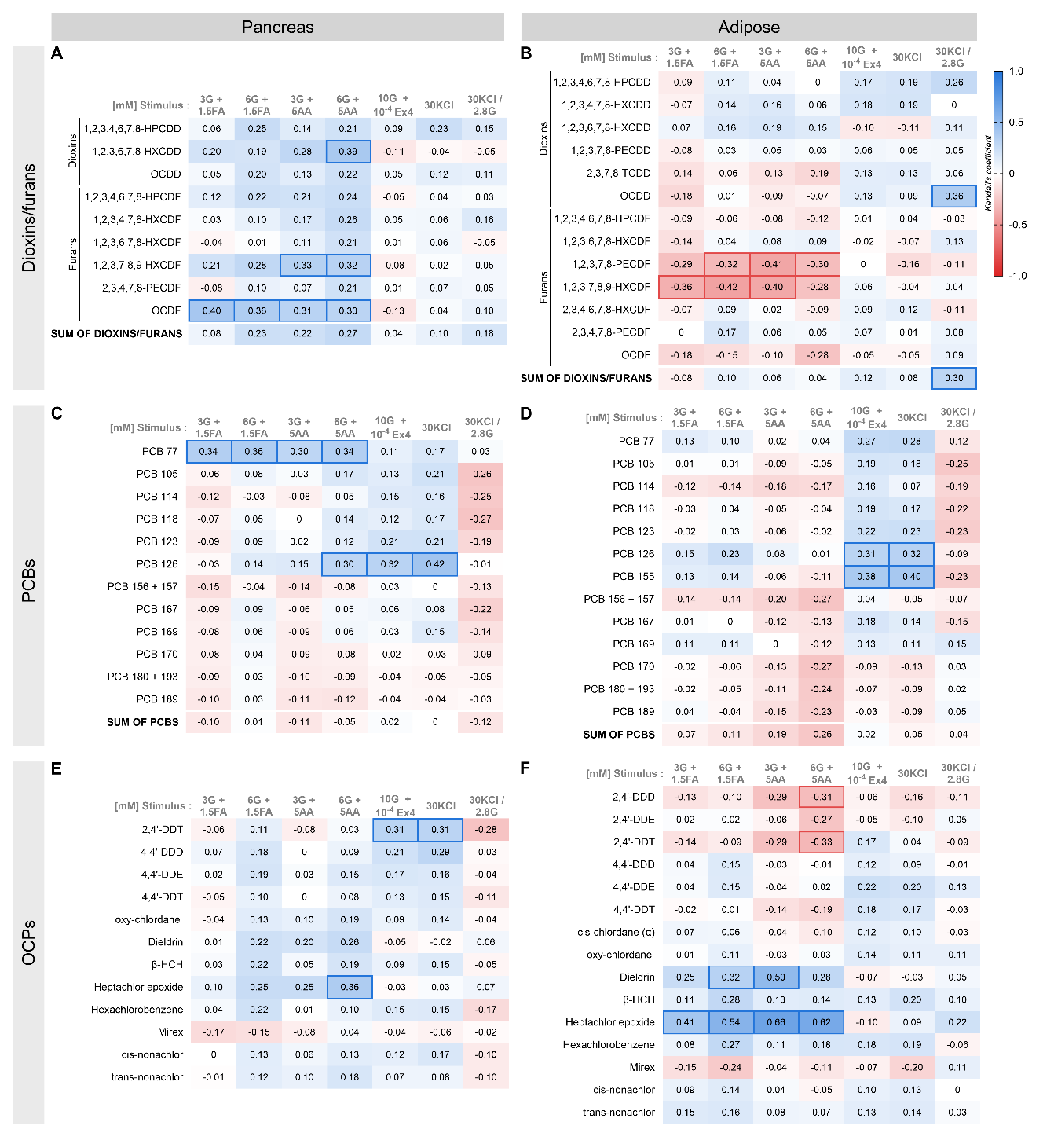

**Supp. Fig. 14: A subset of POPs in pancreas and adipose correlated with fatty acid-, amino acid-, Ex4-, and/or KCl-stimulated insulin secretion.** Heatmaps depicting correlations between **(A,B)** dioxins/furans, **(C,D)** polychlorinated biphenyls (PCBs) and **(E,F)** organochlorine pesticides (OCPs) concentrations in human donor **(A,C,E)** pancreas and **(B,D,F)** adipose and insulin secretion in response to various nutritional stimuli. Human donor islets were stimulated with 3mM glucose (3G) + 1.5mM oleate/palmitate (1:1 mix; 1.5FA), 6mM glucose (6G) + 1.5mM oleate/palmitate, 3mM glucose + 5mM leucine (5AA), 6mM glucose + 5mM leucine, 10mM glucose (10G) + 100 nM Ex4, or 30 mM KCl. Total insulin secretion was measured following each stimulus. Insulin stimulation index in response to KCl was calculated as a ratio of insulin concentration under KCl relative to LG condition. All analyte concentrations used in this analysis were blank corrected and any zeros were assigned a value of 1/2 LOD. Data in the heatmaps represent Kendall’s rank correlation coefficients. Bolded boxes emphasize correlations that are moderate to strong.
